# Associations of common infections with frailty and mortality in two UK cohort studies

**DOI:** 10.1101/2025.11.14.25340163

**Authors:** Demelza Smeeth, Charlotte Warren-Gash, Rebecca E Green, Julia Butt, Tim Waterboer, Alun D Hughes, Nishi Chaturvedi, Dylan M Williams

## Abstract

**Background:** Some common infections are associated with poorer age-related health outcomes; however, findings are limited to a small number of pathogens and frequently inconclusive. This study aimed to expand the range of pathogens investigated in relation to frailty and mortality in older age.

**Methods:** We investigated relationships between seropositivity for 18 viruses, bacteria and protozoa with frailty and mortality in middle-aged and older adults within two UK population based cohorts: UK Biobank (*N*=9,427; aged 40-70 years) and MRC NSHD (*N*=1,791; aged 60-65 years). At baseline, multiplex serological assays were used to identify seropositivity for each pathogen and frailty was assessed using a frailty index which measures the accumulation of age-related health deficits. Mortality was determined from linked administrative records.

**Results:** Previous infection with *Toxoplasma gondii* and *Helicobacter pylori* were associated with higher frailty equivalent to 3.8 or 3.0 years of aging. Inflammation-weighted pathogen burden was also associated with greater frailty. Previous infection with *Chlamydia trachomatis*, human herpes simplex virus 1 and cytomegalovirus were also associated with increased frailty, although relationships were confounded by socioeconomic circumstances. No common infections were robustly associated with mortality.

**Conclusions:** Our results indicate that infection with *H. pylori* and *T. gondii*, and the combined burden of infection may detrimentally impact ageing health. These pathogens may warrant targeting beyond current clinical measures to mitigate the development of frailty.

## Introduction

The global increase in ageing populations has led to an interest in understanding the development and prevention of frailty and other age-related outcomes (1). Frailty reflects an individual’s reduced physiological reserve and increased vulnerability to a wide range of adverse health outcomes, including mortality (2) and can be assessed through the construction of a frailty index (FI) (3,4). This construct uses cumulative counts of age-related health deficits across multiple biological systems, encompassing physical, psychological, biological and social functioning and are associated with a number of negative outcomes including mortality and hospitalisation (5,6). Understanding the factors that contribute to frailty and mortality is essential for developing strategies to promote healthy ageing and reduce healthcare burdens.

Many common pathogens have been linked to health beyond the initial acute infection phase. Some, including those from the *Herpesviridae* family, can establish lifelong latency following primary infection (7). Others, such as *Helicobacter pylori,* may persist chronically and have significant long-term health consequences (8). These infections can adversely affect health directly, through tissue damage and cellular disruption, or indirectly, by promoting systemic inflammation, immune dysregulation, or autoimmunity. Despite the high global prevalence of these persistent infections, the extent to which individual pathogens influence frailty and mortality risk remains poorly understood. Existing studies have explored a narrow range of pathogens and have yielded mixed findings. For example, cytomegalovirus (CMV) seropositivity has been associated with increased frailty and mortality in some populations, but not in others, with differences potentially influenced by sample size, cohort age, health status, and geographic or socioeconomic factors (9–14). Limited numbers of studies have begun to expand the range of pathogens explored, but require replication (15–17).

Cumulative pathogen burden, a composite measure of exposure to multiple pathogens, has also been linked to frailty and mortality, but research remains sparse (17,18). Moreover, the optimal approach to assessing cumulative pathogen exposure requires evaluation. Simple count scores employed to date might have biased any associations of pathogen burden with health towards the null because pathogens with varying pathogenicity are given equal weighting. A more appropriate scoring system would ideally account for differences in pathogenicity of each agent being studied, potentially weighting via their respective inflammatory responses or other underlying shared pathogenic pathways (19).

In the present study we build on existing research and expand the range of pathogens investigated in relation to frailty and mortality in two large population-based studies in older age. We first examined cross-sectional associations of serostatus for 17 individual pathogens with concurrent frailty, measured using FIs. Second, we explored whether serostatus for these pathogens were also associated with incident all-cause mortality risk. Finally, we explored whether the cumulative burden of these pathogens was associated with frailty or mortality.

## Methods

### Cohorts

The UK Biobank (UKB) is large prospective multicentre cohort study with 502,631 participants, aged 40–69 years, enrolled at 22 assessment sites in England, Scotland, and Wales between 2006 and 2010 (20). Baseline assessments captured a wide range of data including demographic, health outcome and biological data. This study used a random subset (n=9,427; 4151 male, 5276 female) whose baseline blood samples were assayed for serology (21). This random subset is representative of the wider UK Biobank cohort.

The Medical Research Council (MRC) 1946 National Survey of Health and Development (NSHD) is a birth cohort study which has followed an initially nationally representative sample of 5,362 individuals since their births in England, Scotland, and Wales in one week in March 1946 (22,23). This research used data on a subset of the cohort (n=1,791; 885 male, 906 female) who participated in the 2006-2010 follow-up at age 60-65 years, which assessed participants’ recent and current health and collected blood samples that were subsequently used for serology assays.

All cohort participants provided written informed consent. Ethical approval for UK Biobank from the National Health Service North-West Research Ethics Committee (11/NW/0382) and for NSHD was obtained from the National Research Ethics Service Committee London (14/LO/1173).

### Multiplex serology and pathogen serostatus

Serum immunoglobulin G (IgG) antibody levels against a selection of antigens from several pathogens (herpesviruses, polyomaviruses, papillomaviruses, bacteria and a protozoan) were measured using a validated fluorescence bead based multiplex serology platform developed at the German Cancer Research Center (DKFZ) in Heidelberg (21,24,25). Antibody responses were quantified as median fluorescence intensity units, and between one and six antigens were quantified per pathogen (eTable 2). In UKB, 21 pathogens were assayed at the baseline assessment. An adaptation of the same multiplex panel assayed 18 pathogens in samples from the NSHD 2006-2010 assessment. Only the 17 pathogens with relevant serology data available in both cohorts were included in this study.

Our primary exposures of interest were serostatus, categorised as binary variables, indicating current and/or past infection for each pathogen (“seropositivity”); these have been previously derived for both UKB and NSHD (see eMethods) (21,26). To measure cumulative exposure to multiple pathogens, we derived a pathogen burden index (PBI) representing the number of positive serostatus values across the 17 pathogens. We also created inflammation-weighted PBIs by weighting each pathogen by its association with either circulating white blood cell count (WBC) or ln-transformed C-reactive protein (CRP) measured from the same blood samples. Regression coefficients for each pathogen were obtained from linear regression models adjusted for age and sex and applied as weights to the corresponding pathogen values and summed to derive the final WBC-weighted PBI for each individual (eTable 3). PBIs were normalised and the z-scores used in all analyses. Similar approaches have been used successfully to explore disease-weighted pathogen burden in the context of stroke and cognitive functioning (27,28).

### Frailty indices

Frailty indices (FIs) have been previously generated for the two cohorts detailed here using data from the baseline UKB questionnaire and self-report items from the age 60-64 NSHD data collection (6,29). For this analysis, these were modified to create new FIs with alignment across the cohorts and following recent guidance on FI creation (30). Items were retained from the existing FIs if they were common to both cohorts, were similarly defined and adhered to the guidelines outlined by Theou and colleagues (no more than 10% missing values, and prevalences greater than 1% and less than 80% in both cohorts) (30). Due to differences in data collected in the two cohorts, some items were adapted to improve consistency. Available data for the two cohorts were screened for further potential FI items. New items had to measure an age- or mortality-related health deficit and adhere to the same rules outlined above. All items were scaled from 0-1, where 0 represents no health deficit and 1 represents the maximum deficit. In total, this resulted in 41 items present in both cohorts, which covered several health domains including pain, cardiometabolic health, infirmity, mental health and respiratory health. The final items were screened to ensure there were no collinear items using Pearson correlation (r<0.95). Further information on FI creation is outlined in the eMethods. FI items were scored for each individual, divided by the maximum score (41) and multiplied by 100 to give a percentage FI with 0% representing no frailty and 100% representing maximum frailty.

### Mortality data

Participants in UKB and NSHD provided consent for their data to be linked to the NHS registers for mortality registrations. Mortality data is available up to August 2024 for NSHD and up to November 2021 for UKB.

### Additional covariates

Cohort participants reported sex and age at assessment. Ethnicity (White, Asian, Black, Mixed, Other) was reported only in UKB; participants in NSHD were exclusively White. In UKB gross household tax before tax was categorised into <£18,000, £18,000-30,999, £31,000-51,999, £52,000-100,000, or >£100,000. Net household income was reported in NSHD and was categorised into <£14,999, £15,000-£29,999, £30,000-£39,999, £40,000-£79,999, or >£80,000. Highest level of education was selected from a list of options in UKB: none, vocational, sub GCE, O-Level, A-Level, degree or higher, or other professional qualifications. Highest level of education in NSHD was partially collapsed in NSHD to yield similar categories to UKB: none, vocational, sub GCE, O-Level, A-Level, or degree or higher.

### Statistical analyses

All analyses were conducted in R (v. 4.4) in RStudio.

Missing covariates and FI items were imputed using multiple imputation by chained equations using the ‘mice’ package in R, using 20 iterations and 20 chains (31). Numerical and binary variables were imputed using predictive mean matching (pmm), ordered factor variables were imputed using the proportional odds model (polr) and unordered factor variables were imputed using polytomous logistic regression (polyreg). Auxiliary variables included body mass index (BMI), smoking status (current/past/never), childhood social class (NSHD only), age at leaving education, employment status, and Townsend deprivation index quintile for their residence at time of interview. Passive imputation was applied to the BMI FI item, PBI, and FI. Convergence was visually assessed through trace-plots, and imputed values were compared with observed data using various diagnostic plots to ensure validity.

Due to differences across studies, analyses were run independently in each individual cohort prior to meta analysis. Analyses in NSHD were conducted using the available sampling weight to account for the socially stratified design (32). All analyses were conducted individually for each pathogen and PBI. All primary descriptive and inferential analyses were conducted across 20 imputed datasets, which were combined using Rubin’s rules. Sensitivity analyses were performed on complete-case datasets to evaluate robustness of findings. Due to the multiple tests performed, findings from meta analyses (per outcome) were corrected for false discovery rate (FDR), using the Benjamini–Hochberg procedure with an alpha of 0.05. However, given that multiple testing correction increases the likelihood of false negative results, suggestive findings (at unadjusted *P* < 0.05) in fully adjusted models are also described.

### Frailty analyses

We used multivariable linear regression models to examine the cross-sectional association of each individual pathogen serostatus or PBI with frailty. All models included the individual pathogen serostatus or PBI as the exposure and FI percentage as the outcome. Minimally adjusted models controlled for participant sex and age at assessment, as well as ethnicity in UKB. Fully adjusted models additionally controlled for income and educational attainment as potential confounders. Outputs are regression betas which represent the adjusted difference in FI percentage in seropositive vs seronegative groups (for individual serostatuses) or per standard deviation increase in PBI with 95% confidence intervals.

The average yearly increase in frailty was calculated to contextualise the results of regression models. This was calculated using data from UKB only due to the limited and skewed age range covered by NSHD. Average yearly increases in frailty were estimated from linear models which modelled the relationship between age at interview and frailty percentage controlling for sex and ethnicity.

### Mortality analyses

We constructed Cox proportional hazard regression models with the ‘survival’ package to examine the association of each individual pathogen and PBI with risk of all-cause mortality (33). The proportional hazards assumption was tested for each model and proportional hazards were assumed where p>0.05. In these models, age at baseline assessment was used as the starting time, and age at censoring or death was used as the time-to-event variable. Models were adjusted as for frailty analyses. Outputs are hazard ratios which represent the adjusted relative hazards of death during follow-up in seropositive vs seronegative groups (for individual serostatuses) or per standard deviation increase in PBI.

### Meta-analysis

Results from the two cohorts were meta-analysed using the ‘metafor’ package (34). The restricted maximum likelihood estimator was used to fit a random-effects model using the rma function. The random-effects model was chosen to account for potential heterogeneity between the two studies, given that variability in study populations or methodologies could influence the results. Regression betas and standard errors were provided for FI analyses and log-hazards, and their accompanying standard errors were provided for survival analyses. Substantial heterogeneity between cohorts was defined as *I* ^2^ > 50% and/or *Q P* value < 0.05. Meta-analysed results for survival analyses were exponentiated to provide hazard ratios for reporting.

## Results

### Cohort descriptions

Full descriptions of the cohorts can be found in Table 1. Both cohorts were balanced for sex (UKB 56.0% female; NSHD 49.4% female) and comprised adults over the age of 40 years (UKB mean age at baseline: 56.5y, SD 8.2; NSHD mean age at 60–64-year follow-up: 63.2y, SD 1.1).

**Table 1.**
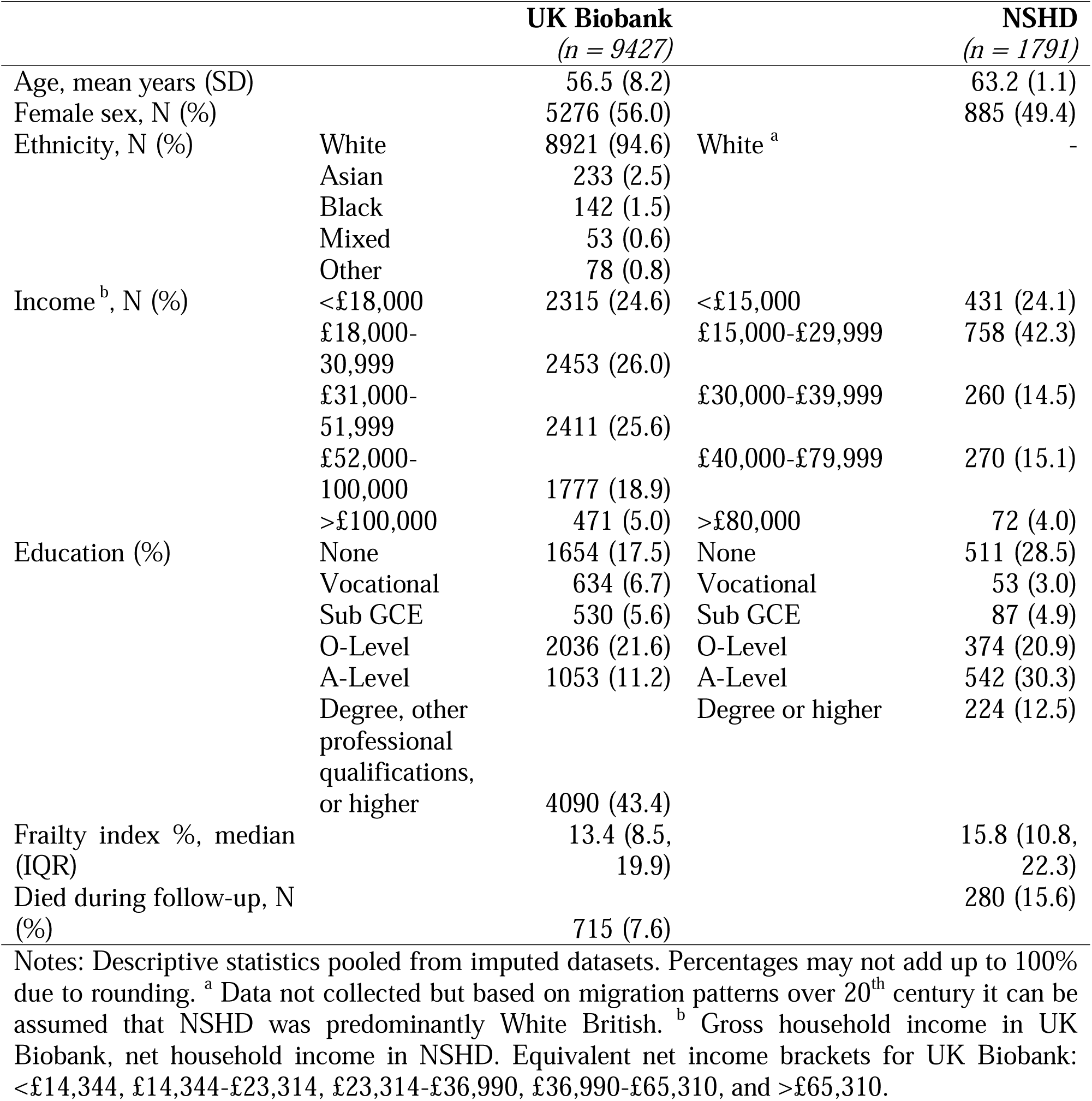
Cohort descriptions for UK Biobank and NSHD.

|  |  | **UK Biobank**<br>(*n* = 9427) |  | **NSHD**<br>(*n* = 1791) |
| --- | --- | --- | --- | --- |
| Age, mean years (SD) |  | 56.5 (8.2) |  | 63.2 (1.1) |
| Female sex, N (%) |  | 5276 (56.0) |  | 885 (49.4) |
| Ethnicity, N (%) | White | 8921 (94.6) | White <sup>a</sup> | - |
|  | Asian | 233 (2.5) |  |  |
|  | Black | 142 (1.5) |  |  |
|  | Mixed | 53 (0.6) |  |  |
|  | Other | 78 (0.8) |  |  |
| Income <sup>b</sup>, N (%) | <£18,000 | 2315 (24.6) | <£15,000 | 431 (24.1) |
|  | £18,000-30,999 | 2453 (26.0) | £15,000-£29,999 | 758 (42.3) |
|  | £31,000-51,999 | 2411 (25.6) | £30,000-£39,999 | 260 (14.5) |
|  | £52,000-100,000 | 1777 (18.9) | £40,000-£79,999 | 270 (15.1) |
|  | >£100,000 | 471 (5.0) | >£80,000 | 72 (4.0) |
| Education (%) | None | 1654 (17.5) | None | 511 (28.5) |
|  | Vocational | 634 (6.7) | Vocational | 53 (3.0) |
|  | Sub GCE | 530 (5.6) | Sub GCE | 87 (4.9) |
|  | O-Level | 2036 (21.6) | O-Level | 374 (20.9) |
|  | A-Level | 1053 (11.2) | A-Level | 542 (30.3) |
|  | Degree, other professional qualifications, or higher | 4090 (43.4) | Degree or higher | 224 (12.5) |
| Frailty index %, median (IQR) |  | 13.4 (8.5, 19.9) |  | 15.8 (10.8, 22.3) |
| Died during follow-up, N (%) |  | 715 (7.6) |  | 280 (15.6) |

Notes: Descriptive statistics pooled from imputed datasets. Percentages may not add up to 100% due to rounding. <sup>a</sup> Data not collected but based on migration patterns over 20<sup>th</sup> century it can be assumed that NSHD was predominantly White British. <sup>b</sup> Gross household income in UK Biobank, net household income in NSHD. Equivalent net income brackets for UK Biobank: <£14,344, £14,344-£23,314, £23,314-£36,990, £36,990-£65,310, and >£65,310.

Seroprevalence of the pathogens studied were generally consistent across the two cohorts, although prevalence for a small number of pathogens was lower in NSHD (Table 2). Most infections were relatively common except for herpes simplex virus 2 (HSV2), Kaposi’s sarcoma-associated herpesvirus (KSHV), and the papillomaviruses (seroprevalences ≤ 16.2%). Accordingly, total pathogen burden was relatively high with most participants testing positive for at least 7 pathogens (UKB mean = 9.0; NSHD mean = 7.4).

**Table 2.** Seroprevalence for the 17 pathogens and pathogen burden indices.

|  |  | **UK Biobank**<br>(*n* = 9427) | **NSHD**<br>(*n* = 1971) |
| --- | --- | --- | --- |
| Herpesviruses, (%) | HSV1 | 69.9 | 67.3 |
|  | HSV2 | 16.2 | 7.3 |
|  | VZV | 92.4 | 79.4 |
|  | EBV | 94.7 | 92.9 |
|  | CMV | 58.3 | 54.2 |
|  | HHV6A | 77.4 | 42.7 |
|  | HHV6B | 79.2 | 53.9 |
|  | HHV7 | 94.7 | 72.8 |
|  | KSHV | 8.1 | 0.3 |
| Papillomaviruses, % | HPV-16 | 4.4 | 2.7 |
|  | HPV-18 | 2.7 | 2.4 |
| Polyomaviruses, % | BK virus | 95.3 | 91.4 |
|  | JC virus | 57.5 | 51.6 |
|  | MC virus | 66.5 | 59.7 |
| Bacteria/protozoa, % | *T. gondii* | 28.0 | 24.4 |
|  | *H. pylori* | 31.7 | 17.9 |
|  | *C. trachomatis* | 21.5 | 17.6 |
| Pathogen burden indices, mean (SD) | Unweighted PBI | 8.98 (2.07) | 7.39 (1.98) |
|  | WBC-weighted PBI | 0.82 (0.24) | 0.28 (0.34) |
|  | CRP-weighted PBI | 0.36 (0.15) | -0.16 (0.10) |

Note: Values pooled from imputed datasets. CMV: cytomegalovirus; CRP: C-reactive protein; EBV: Epstein–Barr virus; HHV: human herpesvirus; HPV: human papillomavirus; HSV: herpes simplex virus; JC: John Cunningham virus; KSHV: Kaposi's sarcoma–associated herpesviruses; MCV: Merkel cell virus; PBI: pathogen burden index; WBC: White blood cells.

FIs were comparable across the cohorts although slightly higher in NSHD as expected given the older mean age of the participants (UKB: median FI: 13.4%, IQR: 8.5, 19.9; NSHD: median: 15.8%, IQR: 10.8, 22.3; Table 1; eFigure 1). Accordingly, many of the individual FI items had higher mean scores in NSHD indicating greater frailty according to that health deficit (eTable 5). FIs were positively correlated with age in UKB *(rs* = 0.19, 95% CI: 0.17, 0.21) and associated with increased risk of mortality in both cohorts (UKB: HR = 1.05 per percentage higher FI, 95% CI: 1.04, 1.06; NSHD: HR = 1.04, 95% CI: 1.03, 1.05) indicating the validity of the measure (30). The FI was not correlated with age in NSHD due to the limited age range covered.

The mean follow-up time was 12.5 years (SD = 0.13) in UKB and 14.3 years (SD = 1.6) in NSHD. Within this study period, 715 (7.6%) UKB participants and 280 (15.6%) NSHD participants had died.

### Pathogen seropositivity and frailty

Meta-analysed results for associations of individual pathogen serostatus with frailty are depicted in Figure 1 and full results are in eTables 6 and 7. Seropositivity for several common infections was associated with higher frailty when compared to seronegative individuals, including *Toxoplasma gondii* (β = 0.90%; 95% CI: 0.55, 1.27)*, H. pylori* (β = 1.26%; 95% CI: 0.90, 1.61), *Chlamydia trachomatis* (β = 0.86%; 95% CI: 0.45, 1.61), human herpes simplex virus 1 (HSV1) (β = 1.19%; 95% CI: 0.58, 1.80), and cytomegalovirus (β = 0.63%; 95% CI: 0.31, 0.96). These point estimates were equivalent to the differences in frailty accrued over 3.1-6.2 years of ageing in UK Biobank. In addition, seropositivity for Epstein–Barr virus and Merkel Cell virus were associated with increased and decreased frailty respectively, but these did not survive controlling for multiple testing. We note that the mean coefficients for EBV’s association with frailty were comparable to that of *H. pylori* and *T. gondii* but were imprecisely estimated due to there being few seronegative individuals in the samples.

**Figure 1.**
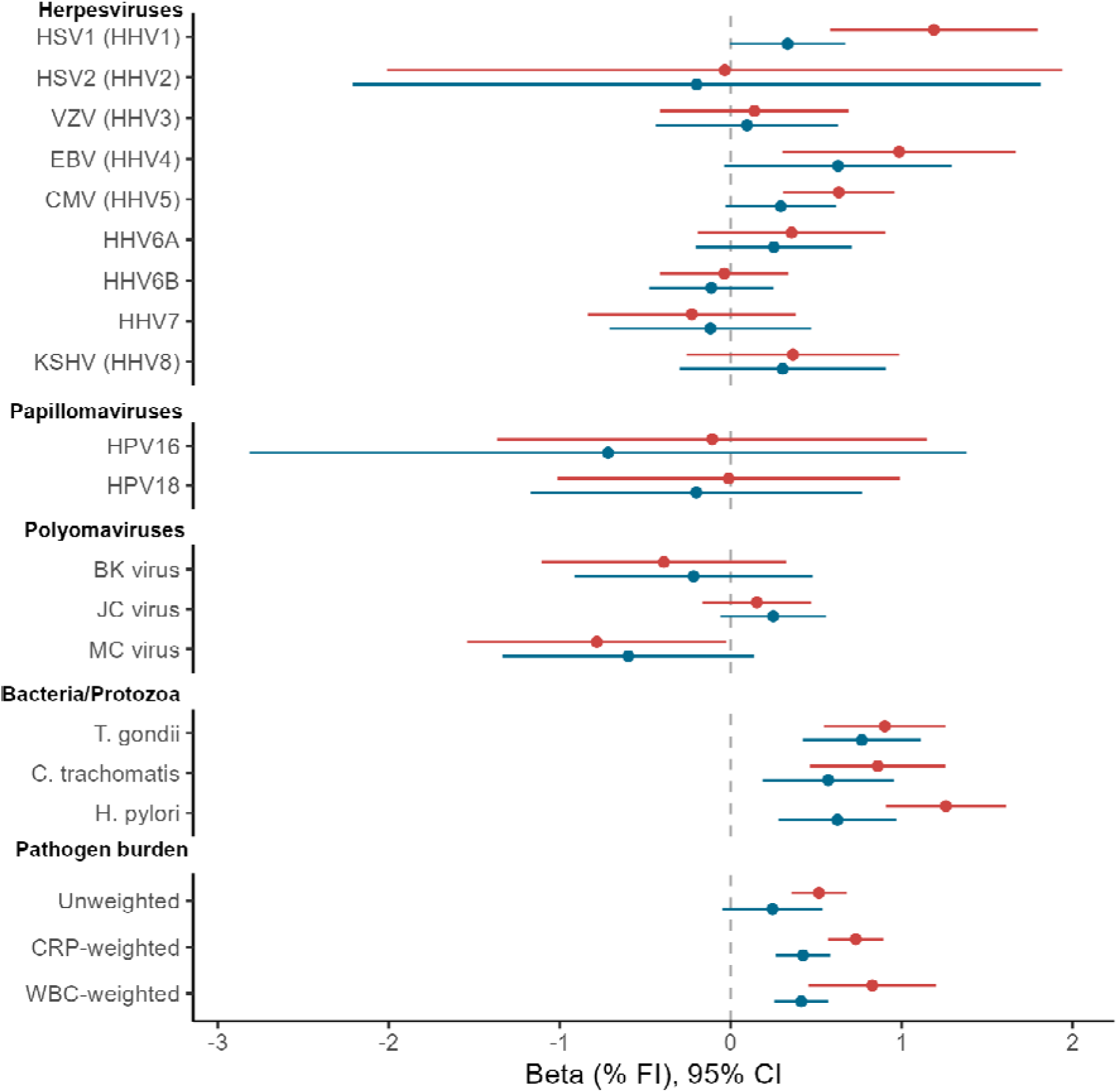
Forest plot indicating meta analysed associations of pathogen serostatus and pathogen burden scores with frailty. Positive estimates indicate increased frailty. Statistical models are indicated by colour. Minimally adjusted models (red) controlled for sex, age, and ethnicity in UKB. Fully adjusted models (blue) additionally controlled for income and educational attainment. Pathogen families are indicated. CI: confidence interval; CMV: cytomegalovirus; EBV: Epstein–Barr virus; HHV: human herpesvirus; HPV: human papillomavirus; HSV: herpe simplex virus; JC: John Cunningham virus; KSHV: Kaposi’s sarcoma–associated herpesviruses; MCV: Merkel cell virus; WBC: White blood cell; CRP: C-reactive protein.

Many of these relationships were attenuated with adjustments for socioeconomic factors. After controlling for income and education, only *Toxoplasma gondii* (β = 0.77%; 95% CI: 0.42, 1.11) and *H. pylori* (β = 0.63%; 95% CI: 0.28, 0.97) were significantly associated with frailty. These coefficients are equivalent to an increase of frailty seen over 3.8 and 3.1 years of ageing respectively. Seropositivity for *Chlamydia trachomatis* was nominally associated with frailty, but this did not survive controls for multiple testing (β = 0.57%; 95% CI: 0.19, 0.95).

In minimally adjusted models, all PBIs were associated with higher frailty, with the inflammation-weighted PBI associations slightly more pronounced (PBI_unwt_: β = 0.52%/SD, 95% CI: 0.36, 0.68; PBI_WBC_: β = 0.83%/SD, 95% CI: 0.45, 1.20; PBI_CRP_: β = 0.73%/SD, 95% CI: 0.57, 0.89). The associations of all three PBIs with frailty attenuated with adjustment for socioeconomic covariates, though statistically robust evidence remained for the associations of inflammation-weighted PBIs (PBI_unwt_: β = 0.25/SD, 95% CI: -0.05, 0.55; PBI_WBC_: β = 0.41%/SD, 95% CI: 0.25, 0.57; PBI_CRP_: β = 0.42%/SD, 95% CI: 0.26, 0.58).

Associations between serostatus variables and frailty were generally consistent in direction between the two cohorts, although effect sizes were generally smaller in NSHD. There was no evidence of heterogeneity where pooled results were statistically significant, but heterogeneity was evident for other pathogens (i.e. where *I^2^* > 50% or *Q* <0.05, eTables 6 and 7). Most notably, HSV2 seropositivity was associated with higher frailty in UKB, but lower frailty in NSHD. For pooled findings, UKB tended to contribute a much greater proportion to the weighting of overall estimates, likely reflecting its larger sample size. Associations estimated from imputed datasets were also generally consistent with complete-case analyses although confidence intervals were substantially narrower, particularly for very rare or common pathogens (eTable 10).

### Pathogen seropositivity and mortality

Compared to frailty, there were fewer clear associations of common pathogens with risk of mortality over a mean follow-up of 13 years (Figure 2; eTables 8 and 9). In minimally adjusted models, only the unweighted PBI was significantly associated with 11% higher risk of mortality after controlling for multiple testing (HR = 1.11, 95% CI: 1.04, 1.18). The WBC-weighted PBI (HR = 1.11, 95% CI: 1.03, 1.19), HSV2 (HR = 1.24, 95% CI: 1.04, 1.47), *T. gondii* (HR = 1.17, 95% CI: 1.02, 1.3) and *C. trachomatis* (HR = 1.19, 95% CI: 1.01, 1.39) were associated with higher mortality risk prior to multiple testing correction while BK virus was associated with a 28% lower risk of mortality (HR = 0.72, 95% CI: 0.57, 0.91).

**Figure 2.**
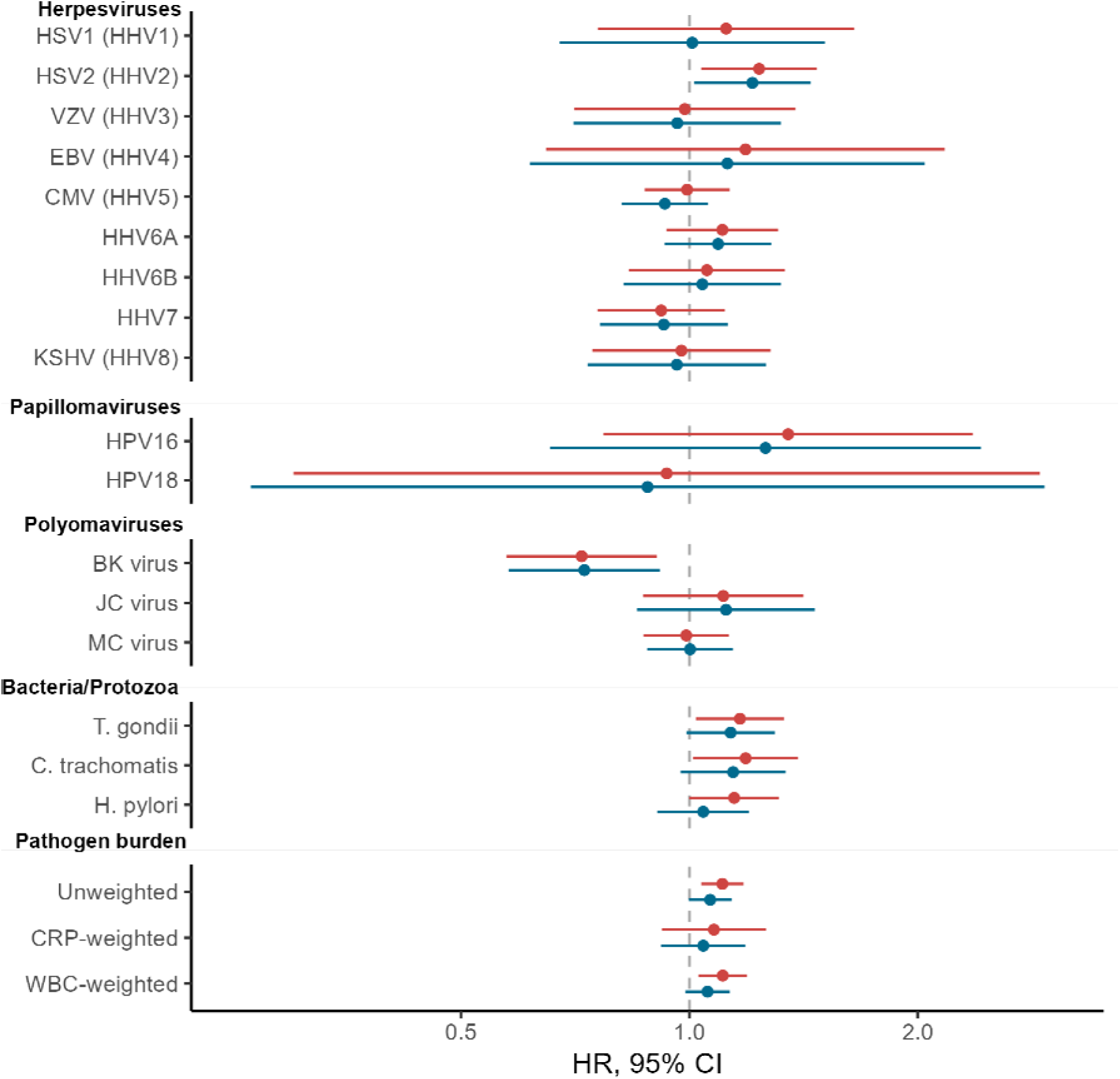
Forest plot indicating meta analysed associations of pathogen serostatus and pathogen burden scores with risk of mortality. Positive estimates indicate increased risk of mortality. Statistical models are indicated by colour. Minimally adjusted models (red) controlled for sex, and ethnicity in UKB. Fully adjusted models (blue) additionally controlled for income and educational attainment. Pathogen families are indicated. CI: confidence interval; CMV: cytomegalovirus; EBV: Epstein–Barr virus; HHV: human herpesvirus; HPV: human papillomavirus; HR: hazard ratio; HSV: herpes simplex virus; JC: John Cunningham virus; KSHV: Kaposi’s sarcoma–associated herpesviruses; MCV: Merkel cell virus; WBC: White blood cell; CRP: C-reactive protein.

Following adjustment for socioeconomic factors and correction for multiple testing, no serology measures were significantly associated with mortality. However, HSV2 seropositivity was nominally associated with an elevated higher risk of mortality (HR = 1.21, 95% CI: 1.01, 1.44), while BK virus seropositivity was nominally associated with a lower risk of mortality (HR = 0.73, 95% CI: 0.58, 0.91). Associations from imputed datasets were generally consistent with complete-case analyses.

## Discussion

In this cross cohort study using two British population based samples of middle-aged to older adults, we found that exposure to several common infections were associated with increased frailty, although only two were independent of individuals’ socioeconomic circumstances. Accordingly, we also found that a higher cumulative pathogen burden was associated with greater frailty, however this only survived controlling for socioeconomic factors when the pathogen burden index was weighted by the degree to which pathogens associated with systemic inflammation. We did not find evidence that individual pathogens or their combined burden were associated with mortality during the follow-up period of 10-15 years, though there were several suggestive findings that would warrant further examination.

The clearest evidence of individual pathogen associations with frailty was observed for the two bacteria (*H. pylori and C. trachomatis)* and protozoan (*T. gondii*). To our knowledge, this is the first investigation of *H. pylori* and *C. trachomatis* exposure in relation to frailty, although *H. pylori* infection has been linked to multi-system morbidity and mortality (35). *C. trachomatis* and *H. pylori* are responsible for some of the most frequent chronic infections in humans. Despite their prevalence, symptomatic infections are scarce, but these pathogens are associated with elevated peripheral inflammation which has been hypothesised to lead to more systemic impacts in addition to known clinical consequences, such as a role of *H. pylori* in peptic ulcer disease (19). The protozoal infection, *T. gondii,* has been previously linked with increased frailty in one small case-control study, which did not find an association of seropositivity with frailty, but found a positive association between seroreactivity for *T. gondii* and frailty among seropositive individuals (16). *T. gondii* is also quite prevalent (seroprevalences >20% in both current cohorts) and can establish latent and often asymptomatic infections. While more commonly associated with neuropsychological outcomes (36), latent *T. gondii* infection has also been associated with elevated allostatic load, a factor involved in the development of frailty (37).

Previous research on the association between pathogen serostatus and frailty has tended to focus predominantly on herpesviruses, and particularly CMV given its relationship to the ageing immune system (38). Such studies suggest that CMV may be associated with increased frailty, but this is limited to those aged 60-75 years and associations are inconsistent (17,39,40). In this study, while CMV was associated with higher frailty in minimally adjusted models, this relationship attenuated with adjustment for socioeconomic covariates (particularly in NSHD), potentially explaining the inconsistent findings across different populations or settings. Similar changes to associations were also found for HSV1 and EBV, which alongside existing research on other herpesviruses, suggest that these associations might be confounded by socioeconomic factors (17,38). However, some caution is needed regarding the interpretation of models adjusted for socioeconomic factors: whilst these factors could plausibly confound associations, it is also possible that infections may mediate some of the effects of education and other socioeconomic conditions on these health outcomes.

Despite several associations of pathogen serostatus with frailty, there was limited evidence for associations with mortality over the study period. However, results for HSV2, *T. gondii and C. trachomatis* were all at least suggestive of higher mortality risk among seropositive individuals, while BK virus was suggestive of reduced mortality risk among seropositive individuals. Seropositivity for three of the herpesviruses and *H. pylori* has previously been tested for associations with all-cause mortality in UKB (15,35), but our analyses incorporate more pathogens, more recent mortality data and are included here for completeness for meta-analysing alongside NSHD data. The potential relationships of *T. gondii* and *C. trachomatis* with mortality risk are made more plausible when considered in parallel to their clearer associations with frailty and are also supported by some limited existing research (41). HSV2 was not associated with frailty in the meta-analysed findings due to heterogenous associations across the two cohorts, but has been previously associated with both frailty and mortality in women (14). A number of studies have found reduced survival rates in seropositive individuals, particularly for CMV as well as *H. pylori* (10,11,42,43), although other studies have found null relationships for *H. pylori* or other herpesviruses (14,35,44). As noted previously, studies that detected a positive association between CMV serostatus and mortality often have larger sample sizes or a greater number of mortality events than in this study (15). It is not clear why BK virus was associated with increased survival, but given infection is common in childhood, it is possible that this relationship has been shaped over the long lag time between initial infection and the study period. The meta-analysis of findings between the current two cohorts may also have masked some specific findings of note: for instance, HPV16 and EBV had the strongest associations with mortality risk in UKB, but neither association was present in NSHD. If these were not chance findings in UKB, it may be that these pathogens are associated with mortality at younger ages (middle age) as opposed to older age as studied in NSHD.

In addition to assessing individual pathogens, we also considered the graded relationships with total pathogen burden i.e., the total number of pathogens that an individual tested positive for. Total pathogen burden was associated with increased risk of frailty and mortality in minimally adjusted models but there were degrees of attenuation when adjustments for socioeconomic factors were added. Multiple previous studies have found a positive association between pathogen burden and mortality (17,43,45) ^17,29,31^ and while not investigated in relation to frailty directly before now, pathogen burden was associated with frailty-related biomarkers (18). In these studies, all pathogens were given equal weighting, which risks underestimating the impact of pathogens detrimental to health by giving them equal weighting to those that have little or no pathogenic effect. This is especially true for this study due to the greater number of pathogens studied. To address this concern, we created inflammation-weighted PBIs. There were some differences in the results for the unweighted and weighted PBIs - the inflammation-weighted PBIs had significantly stronger relationships with frailty, potentially reflecting the relative importance of inflammation in ageing, though unweighted and weighted PBIs did not differ clearly in terms of their associations with mortality.

This study has several strengths. It is the largest to date in terms of number of pathogens examined (17 agents) and included a large, combined sample of 11,398 total participants. This allowed us to detect novel relationships between seropositivity and frailty. We used data from two large and well characterised population based cohorts which allowed the creation of a detailed FI encompassing a wide range of physiological domains with similar items across the two cohorts. Such indices have been associated with a number of relevant outcomes such as mortality and hospitalisation (5,6). The use of multiplex serology data to directly ascertain pathogen exposure avoids the potential inaccuracies associated with either self-report of past infection, and avoids relying on diagnosis of potentially asymptomatic infections from linked electronic health records. Finally, the use of linked mortality records ensured accurate mortality data over a long follow-up period.

Several limitations require discussion. First, while the cohorts are community samples, they are not representative of the UK population. Participants remaining in NSHD were only broadly representative of the study population at recruitment and selective attrition will have further biased the sample (22). Participants in UKB are healthier and more socially advantaged than the UK population (46), however associations between risk factors and mortality are similar to more representative samples (47). Secondly, analyses are observational and, in the case of the frailty analyses, cross-sectional. Consequently, associations may arise from reverse causality, residual confounding, or other sources of unmeasured bias. Therefore, causation cannot be established without longitudinal studies, particularly as there is evidence that frailty may increase the risk of recent infection (48). Third, despite our large sample size, some findings lacked precision where seropositivity for pathogens was either very high or very low. Alongside the somewhat limited number of mortality events, we chose not to examine associations of seroreactivity levels for antigens with each outcome, for which testing may have suffered from limited statistical power. Fourth, serostatus for this set of pathogens was only assessed once and individuals may have seroconverted in either direction either before or after serological assessment (49). We were unable to retrospectively identify the age or severity of infection, limiting investigation of these factors in relation to the outcomes. Finally, we cannot rule out the potential impact of other non-measured pathogens, especially those that frequently co-occur with those measured here.

In conclusion, our findings add to evidence suggesting that exposure to multiple pathogens may be detrimental to lifelong health, even in the absence of symptomatic disease. We find that *H. pylori* and *T. gondii* may warrant targeting beyond their immediate acute infections and other pathogens such as *C. trachomatis*, HSV1, EBV and CMV may also merit further investigation. Examining these associations in larger datasets as they become available will help to establish or refute some of the more marginal associations we observed and enable the testing of the relevance of seroreactivity (recent activity against pathogens) for these outcomes. Given that seropositivity for several pathogens appears to be a risk factor for frailty and/or mortality risk, future research should also focus on assessing whether or not these associations represent causation, e.g. through other study designs that aid causal inference, such as Mendelian randomisation (50). This would help to motivate whether preventative measures or treatments for relevant pathogens could be tested as interventions to promote healthy ageing.

## Contributions

Conceptualisation: DS, CWG, TW, ADH, NC, DMW; Data curation: DS, REG, DMW; Formal analysis: DS; REG, DMW; Investigation: DS, REG, JB, TW, DMW; Methodology: DS, CWG, REG, JB, TW, ADH, NC, DMW; Project administration: DS, NC, DMW; Supervision: NC, DMW; Visualization: DS; Writing – original draft: DS, DMW; Writing – review and editing: DS, CWG, REG, JB, TW, ADH, NC, DMW.

## Data Availability

This research has been conducted using the UK Biobank Resource under application number 71702. UK Biobank data can be accessed through the UK Biobank Research Analysis Portal (https://www.ukbiobank.ac.uk/enable-your-research). This work uses data provided by patients and collected by the NHS as part of their care and support. Re-used with the permission of the NHS England and UK Biobank. All rights reserved. We are extremely grateful to the NSHD study members for their lifelong participation and continuing support; and to past and present members of the study teams, who helped to collect and process the data. NSHD data used in this publication are available to bona fide researchers upon request to the NSHD Data Sharing Committee via a standard application procedure. Further details can be found at http://www.nshd.mrc.ac.uk/data; doi: 10.5522/NSHD/Q101; doi: 10.5522/NSHD/Q102.

## Acknowledgments

The contributing studies have been made possible because of the tireless dedication, commitment and enthusiasm of the many people who have taken part. We would like to thank the participants and the numerous team members involved in the studies including interviewers, technicians, researchers, administrators, managers, health professionals and volunteers. We are additionally grateful to our funders for their financial input and support in making this research happen. This research has been conducted using the UK Biobank Resource under application number 71702. UK Biobank data can be accessed through the UK Biobank Research Analysis Portal (https://www.ukbiobank.ac.uk/enable-your-research). This work uses data provided by patients and collected by the NHS as part of their care and support. Copyright © 2023, NHS England. Re-used with the permission of the NHS England and UK Biobank. All rights reserved. We are extremely grateful to the NSHD study members for their lifelong participation and continuing support; and to past and present members of the study teams, who helped to collect and process the data. NSHD data used in this publication are available to bona fide researchers upon request to the NSHD Data Sharing Committee via a standard application procedure. Further details can be found at http://www.nshd.mrc.ac.uk/data; doi: 10.5522/NSHD/Q101; doi: 10.5522/NSHD/Q102.

