## Supplemental information for "Associations of common infections with frailty and mortality in two UK cohort studies"

### eMethods

#### Serology measures

Serology measures were derived from blood serum samples collected at baseline assessment in the UK Biobank (UKB) and the age 60-64 assessment in the MRC National Survey of Health and Development (NSHD). At baseline, 10,110 UK Biobank participant samples were selected at random and assayed at German Cancer Research Centre in Heidelberg, Germany in July 2016 using multiplex serology. Of these, 9,427 samples passed various quality control checks and are available for data analysis. 1,813 NSHD participant samples were assayed at German Cancer Research Centre in Heidelberg, Germany in October 2022 using an adaptation of the same methodology. Of these, 1,791 passed quality control checks and are available for data analysis. Technical failures in the assaying included pipetting errors, high background, and insufficient beadcounts. Other exclusions included withdrawal of consent for use of data. Antibody responses for each pathogen were quantified in median fluorescence intensity units (MFU), and between one and six antigens were quantified per pathogen (eTable 2).

We used serostatus variables provided by UKB, and derived serostatus in NSHD using cut-offs recommended by the developers of the multiplex serology platform (Supplementary Table 2). UKB variables were extracted using field codes and serostatus derivation in NSHD has been described previously (1). In UKB, 21 pathogens were assayed at the baselines assessment. An adaptation of the same multiplex panel assayed 18 pathogens at the NSHD 2006-2010 wave. In addition to the listed pathogens, data was also available for human immunodeficiency virus-1 (HIV-1), hepatitis B, hepatitis C, and human T-cell leukemia virus-1 (HTLV-1) in UKB, and for *F.nucleatum* in NSHD. As analyses were restricted to the 17 pathogens common to both studies, these pathogens have not been further described.

Pathogen burden indices (PBIs) were calculated by summing the number of positive serostatus values across the 17 pathogens. We also created inflammation-weighted PBIs which gave greater emphasis to long-term pathogens which were associated with greater systemic inflammation. Analyses were conducted using C-reactive protein (CRP) concentration or white blood cell (WBC) count weighted PBIs derived using blood CRP concentrations or WBC counts collected at the same time. Linear regression models were fitted with either log-transformed CRP concentration or WBC count as the outcome, serostatus for each pathogen as the primary exposure, and were adjusted for sex and age. Regressions were performed independently for each cohort, pathogen serostatus, and inflammation measure. Regression coefficients were extracted and applied as weights to the corresponding pathogen values and summed to derive the final CRP or WBC-weighted PBIs for each individual per cohort. Derived weights are found in eTable 3.

#### Frailty index

Existing frailty indices (FIs) developed for UKB (2) and NSHD at the age 60-64 data collection wave (3) were reviewed for cross-cohort applicability. Items from the UKB FI were dropped if there was no equivalent data available in NSHD (dental problems, misery, loneliness, neck pain, stomach pain, hip pain, facial pain, hiatus hernia, diverticulitis, and migraine). Items were also dropped if the outcome was too rare in either cohort (<1%; psoriasis, multiple cancers, deep vein thrombosis and gall stones). Items from the NSHD FI were dropped if there was no equivalent data in UKB (heart rhythm abnormality, language and speech deficits, walking difficulties, upper limb dysfunction, restriction of physical activities, shingles), it was too rare in either cohort (<1%; heart failure, other cardiovascular disease, other lung conditions, other musculoskeletal problems, other mental health problems), there was too much missing data in either cohort (>10%; cognitive dysfunction, liver diseases, kidney diseases) or it didn’t represent an age-related health deficit (smoking, lack of immunisations, congenital heart disease, rheumatic heart disease).

Some items were adapted so that definitions were better aligned across the two cohorts. The asthma and allergy UKB items were combined, the back pain and sciatica UKB items were combined, head pain was separated from neck pain in the UKB item, occurrence of bronchitis was added to the pneumonia UKB item, indigestion was added to the GERD UKB item, and the hypothyroidism UKB FI item was expanded to include any thyroid problem.

The available data from UKB and NSHD were reviewed for additional health deficits that could be applied to both cohorts. Unhealthy body mass index (BMI), waist-to-hip ratio, chronic bronchitis, polypharmacy and recent weight loss were added. A number of other items were considered but were not retained due to scarcity or excessive missing data. The final list of items and their definitions is available in eTable 4.

#### Imputation

The imputation method we used – multiple imputation by chained equations – assumes data are missing at random. To understand possible sources of missingness, we assessed the quantity of missing data and visualised missingness patterns. We found no evidence to suggest that data were not missing at random. Missing covariates and FI items (serostatuses were complete) were imputed using multiple imputation by chained equations using the mice package in R, using 20 iterations and 20 chains (4). Auxiliary variables were selected based on their prediction of missingness or variables with missing data. Auxiliary variables included BMI, smoking status (current/past/never), childhood social class (NSHD only), age at leaving education, employment status, and Townsend deprivation index quintile for participant residence at time of interview.

Numerical and binary variables were imputed using predictive mean matching (pmm), ordered factor variables were imputed using the proportional odds model (polr) and unordered factor variables were imputed using polytomous logistic regression (polyreg). Passive imputation was applied to the BMI FI item, PBI, and FI variables. Convergence was visually assessed through traceplots, and imputed values were compared with observed data using various diagnostic plots to ensure plausibility. Descriptive statistics for the complete case and multiple imputed datasets can be found in eTable 1.

#### Follow-up analyses

Complete-case analyses were carried out on datasets where cases with missing FI or covariate data were removed. FIs were calculated where less than 10% of FI items were missing, therefore frailty analyses were repeated on imputed datasets excluding those with at least 10% of FI items missing (data not shown). As frailty indices were left skewed (eFigure 1), we also repeated frailty analyses with square root-transformed values to ensure that we were not missing non-linear relationships (data not shown). All sensitivity analyses were conducted across both cohorts and meta-analysed as in the primary analyses.

### eFigures

eFigure 1. Histograms depicting frailty indices across the UK Biobank and NSHD cohorts.
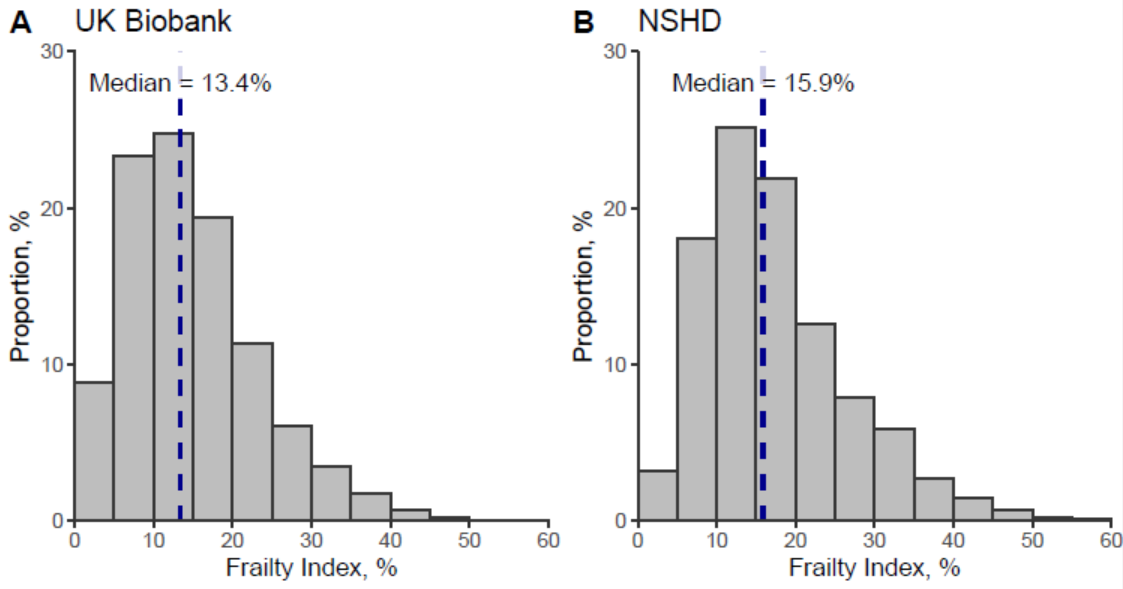

### eTables

eTable 1. Cohort descriptions for complete and multiple imputed datasets

|  | **UKB** | | | **NSHD** | | |
| --- | --- | --- | --- | --- | --- | --- |
|  |  | Complete case | Multiple imputed |  | Complete case | Multiple imputed |
| N |  | 6216 | 9427 |  | 1262 | 1791 |
| Age, mean years (SD) |  | 56.2 (8.2) | 56.5 (8.2) |  | 63.1 (1.1) | 63.2 (1.1) |
| Female sex, N (%) |  | 2819 (45.4%) | 5276 (56.0) |  | 634 (50.2) | 885 (49.4) |
| Ethnicity, N (%) | White | 5957 (95.8%) | 8921 (94.6) | White^a^ | ~100% | ~100% |
|  | Asian | 113 (1.8%) | 233 (2.5) |  |  |  |
|  | Black | 73 (1.2%) | 142 (1.5) |  |  |  |
|  | Mixed | 31 (0.5%) | 53 (0.6) |  |  |  |
|  | Other | 42 (0.7%) | 78 (0.8) |  |  |  |
| Income, N (%) | <£18,000 | 1329 (21.4%) | 2315 (24.6) | <£14,999 | 289 (22.9) | 431 (24.1) |
|  | £18,000-30,999 | 1614 (26%) | 2453 (26.0) | £15,000-£29,999 | 553 (43.8) | 758 (42.3) |
|  | £31,000-51,999 | 1650 (26.5%) | 2411 (25.6) | £30,000-£39,999 | 173 (13.7) | 260 (14.5) |
|  | £52,000-100,000 | 1260 (20.3%) | 1777 (18.9) | £40,000-£79,999 | 201 (15.9) | 270 (15.1) |
|  | >£100,000 | 363 (5.8%) | 471 (5.0) | >£80,000 | 46 (3.6) | 72 (4.0) |
| Education (%) | None | 2225 (35.8%) | 1654 (17.5) | None | 334 (26.5) | 511 (28.5) |
|  | Vocational | 883 (14.2%) | 634 (6.7) | Vocational | 32 (2.5) | 53 (3.0) |
|  | Sub GCE | 728 (11.7%) | 530 (5.6) | Sub GCE | 56 (4.4) | 87 (4.9) |
|  | O-Level | 324 (5.2%) | 2036 (21.6) | O-Level | 279 (22.1) | 374 (20.9) |
|  | A-Level | 1353 (21.8%) | 1053 (11.2) | A-Level | 383 (30.3) | 542 (30.3) |
|  | Degree or higher | 320 (5.1%) | 3037 (32.2) | Degree or higher | 178 (14.1) | 224 (12.5) |
|  | Other professional qualifications | 383 (6.2%) | 484 (5.1) |  |  |  |
| Frailty index %, median (IQR) |  | 13.0 (8.1, 18.7) | 13.4 (8.5, 19.9) |  | 15.2 (10.8, 21.7) | 15.8 (10.8, 22.3) |
| Died during follow-up, N (%) |  | 457 (7.4) | 715 (7.6) |  | 183 (14.5) | 280 (15.6) |
| Unweighted PBI, mean (SD) |  | 8.9 (2.1) | 9.0 (2.1) |  | 7.4 (2.0) | 7.4 (2.0) |
| Notes: Percentages may not add up to 100% due to rounding. ^a^ Data not given as NSHD was predominantly White British. | | | | | | |

eTable 2. Definition of pathogen serostatuses

|  | **UKB** | **NSHD** |
| --- | --- | --- |
| **Herpesviruses** |  |  |
| Herpes simplex virus-1 | 1gG; cut-off: 150; [23050] | 1gG; cut-off: 170 |
| Herpes simplex virus-2 | 2mG; cut-off: 150 [23051] | 2mgG unique; cut-off: 180 |
| Varicella zoster virus | gE / gI; cut-off: 100 [23052] | gE / gI; cut-off: 100 |
| Epstein-Barr virus | ≥2 positive out of EBNA-1 (cut-off: 250), EA-D (cut-off: 100), VCAp18 (cut-off: 250), ZEBRA (cut-off: 100) [23053] | ≥2 positive out of EBNA-1 (cut-off: 411), EA-D (cut-off: 110), VCAp18 (cut-off: 2526), ZEBRA (cut-off: 74) |
| Cytomegalovirus | ≥2 positive out of pp150N (cut-off: 100), pp52 (cut-off: 150) and pp28 (cut-off: 200) [23054] | ≥2 positive out of pp150N (cut-off: 100), pp52 (cut-off: 854), pp28 (cut-off: 100) |
| Human herpesvirus-6A | IE1A; cutoff: 100 [23056] | IE1A (cut-off: 100) AND/OR p100 (cut-off: 75) |
| Human herpesvirus-6B | IE1B; cut-off: 100 [23057] | IE1B (cut-off: 100) AND/OR p101K (cut-off: 100) |
| Human herpesvirus 7 | U14; cut-off: 100 [23058] | U14; cut-off: 225 |
| Kaposi’s sarcoma-associated virus | LANA (cut-off: 100) AND/OR K8.1 (cut-off: 175) [23059] | LANA 3 (100) AND K8.1 (100) |
| **Polyomaviruses** |  |  |
| BK virus | VP; cut-off: 250 [23065] | VP1; cut-off: 250 |
| JC virus | VP1; cut-off: 250 [23066] | VP1; cut-off: 100 |
| Merkel Cell virus | VP1; cut-off: 250 [23067] | VP1; cut-off: 250 |
| **Papillomaviruses** |  |  |
| Human Papillomavirus-16 | L1: cut-off: 175 [23068] | L1; cut-off: 100 |
| Human Papillomavirus-18 | L1: cut-off: 175 [23069] | L1; cut-off: 100 |
| **Bacteria/protozoa** |  |  |
| *T.gondii* | p22 (cut-off: 100) AND/OR sag1 (cut-off: 160) [23062] | p22 (cut-off: 150) AND/OR sag1 (cut-off: 150) |
| *H.pylori* | ≥2 positive out of 5:  VacA (cut-off: 100), OMP (cut-off: 170), GroEL (cut-off: 80), Catalase (cut-off: 180) and UreA (cut-off: 130) [23074] | ≥3 positive out of 8:  GroEL (cut-off: 100); Hcp C (cut-off: 200); HP 1564 (cut-off: 400); VacA-C (cut-off: 250); CagA-N (cut-off: 400); HP305 (cut-off: 150); UreA (cut-off: 250); Catalase (cut-off: 400) |
| *C.trachomatis* | pGP3; cut-off: 200 [23070] | pGP3; cut-off: 95 |
| Notes: Square bracketed numbers for UKB antigens represent the UKB field code for seropositivity. Cut-offs given in MFIs. | | |

eTable 3. Weights for inflammation-weighted pathogen burden indices

|  | **WBC** | | **CRP** | |
| --- | --- | --- | --- | --- |
|  | UKB | NSHD | UKB | NSHD |
| **Herpesviruses** |  |  |  |  |
| Herpes simplex virus-1 | 0.193 | 0.364 | 0.094 | 0.076 |
| Herpes simplex virus-2 | -0.069 | -0.192 | 0.041 | -0.061 |
| Varicella zoster virus | 0.078 | -0.069 | 0.051 | 0.014 |
| Epstein-Barr virus | 0.263 | 0.101 | 0.048 | 0.063 |
| Cytomegalovirus | 0.066 | 0.073 | 0.092 | 0.007 |
| Human herpesvirus-6A | 0.176 | 0.131 | 0.094 | 0.006 |
| Human herpesvirus-6B | 0.051 | 0.089 | 0.015 | -0.027 |
| Human herpesvirus 7 | 0.036 | 0.009 | 0.040 | -0.097 |
| Kaposi’s sarcoma-associated virus | 0.002 | -0.371 | 0.068 | -0.333 |
| **Polyomaviruses** |  |  |  |  |
| BK virus | 0.005 | -0.063 | -0.044 | -0.158 |
| JC virus | -0.002 | -0.034 | -0.001 | -0.002 |
| Merkel Cell virus | -0.009 | -0.212 | -0.042 | -0.054 |
| **Papillomaviruses** |  |  |  |  |
| Human Papillomavirus-16 | 0.08 | -0.354 | 0.091 | -0.014 |
| Human Papillomavirus-18 | -0.032 | -0.622 | 0.185 | -0.136 |
| **Bacteria/protozoa** |  |  |  |  |
| *T.gondii* | 0.198 | -0.109 | 0.091 | -0.078 |
| *H.pylori* | 0.197 | 0.346 | 0.105 | 0.038 |
| *C.trachomatis* | 0.044 | 0.284 | 0.057 | -0.045 |
| Notes: BK: BK virus; CMV: cytomegalovirus; EBV: Epstein–Barr virus; HHV: human herpesvirus; HPV: human papillomavirus; HSV: herpes simplex virus; JC: John Cunningham virus; KSHV: Kaposi's sarcoma–associated herpesviruses; MCV: Merkel cell virus; CRP: C-reactive protein; WBC: white blood cell. | | | | |

eTable 4. Definitions of frailty index items

| **Item** | **NSHD definition** | **UKB definition** | **Scoring** |
| --- | --- | --- | --- |
| Cancer | Self-report of ever having had cancer OR cancer flagged on the cancer registry | Self-report of ever having had cancer diagnosed | No = 0; yes = 1 |
| Anaemia | Currently taking medications for anaemia (drugs that replace iron in iron deficiency anaemia or treat megaloblastic or hypoplastic anaemias) OR haemoglobin <13 g/dl (males) or haemoglobin <12 g/dl (females) ^5^ | Self-report of ever having had anaemia diagnosed (including iron deficiency, aplastic, pernicious) | No = 0; yes = 1 |
| Angina | Self-report of ever having had angina diagnosed | Self-report of ever having had angina diagnosed | No = 0; yes = 1 |
| Chest pain | Self-report of ever having had pain or discomfort in their chest | Self-report of ever having had pain or discomfort in their chest | No = 0; yes = 1 |
| Diabetes | Self-report of ever having had diabetes diagnosed OR use of diabetic medications (inc. insulin) | Self-report of ever having had diabetes diagnosed | No = 0; yes = 1 |
| Myocardial infarction | Self-report of ever having had myocardial infarction | Self-report of ever having had myocardial infarction | No = 0; yes = 1 |
| High blood pressure | Self-report of ever having had high blood pressure diagnosed OR elevated blood pressure readings (systolic >140 mmHg or diastolic >80mmHg) OR use of blood pressure medication | Self-report of ever having had high blood pressure diagnosed OR elevated blood pressure readings (systolic >140 mmHg or diastolic >80mmHg) OR use of blood pressure medication | No = 0; yes = 1 |
| Raised cholesterol | Use of lipid-lowering medications OR total:HDL cholesterol ratio ≥6.0mmol/l | Use of lipid-lowering medications OR total:HDL cholesterol ratio ≥6.0mmol/l | No = 0; yes = 1 |
| Stroke | Self-report of ever having had stroke | Self-report of ever having had stroke | No = 0; yes = 1 |
| Thyroid disorder | Self-report of ever having had a thyroid disorder OR use of thyroid medications | Self-report of ever having had a thyroid problems, hypothyroidism or hyperthyroidism | No = 0; yes = 1 |
| BMI | Underweight: BMI ≤ 18.5; overweight: 25 ≥ BMI < 30; obese: BMI ≥ 30 | Underweight: BMI ≤ 18.5; overweight: 25 ≥ BMI < 30; obese: BMI ≥ 30 | Healthy = 0; overweight = 0.5; underweight or obese = 1 |
| Waist to hip ratio | Males: WHR ≥ 1; Females: WHR ≥ 0.85 | Males: WHR ≥ 1; Females: WHR ≥ 0.85 | No = 0; yes = 1 |
| Recent weight lost | Self-report of unexplained weight loss over the past 12 months | Self-report of weight loss compared to one year ago | No = 0; yes = 1 |
| Epilepsy | Self-report of ever having had a epilepsy OR use of epileptic medications | Self-report of ever having had a epilepsy | No = 0; yes = 1 |
| General health | Self-rated general health on a 5-point scale (excellent very good, good, fair, poor) | Self-rated general health on a 4-point scale (excellent, good, fair, poor) | Scaled excellent to poor on scale of 0-1 (NSHD: 0, 0.25, 0.5, 0.75, 1; UKB: 0, 1/3, 2/3, 1) |
| Polypharmacy | Self-report of currently taking at least 5 medications | Self-report of currently taking at least 5 medications | No = 0; yes = 1 |
| Long-term illness | Self-report of long-term illness, health problem or disability that limits the activities/work they can do | Self-report of any long-standing illness, disability or infirmity | No = 0; yes = 1 |
| Falls | Self-report of having fallen in the past 12 months | Self-report of at least one fall in the past year | No = 0; yes = 1 |
| Fractures | Self-report of having broken a bone in the past 5 years | Self-report of having broken a bone in the past 5 years | No = 0; yes = 1 |
| Anxiety | Frequency of feeling nervous and strung up all the time in the past 2 weeks compared to usual (not at all, no more than usual, rather more than usual, much more than usual) | Frequency of feeling tense, fidgety or restless in the past 2 weeks (not at all, several days, more than half the days, nearly every day) | Scaled on scale of 0-1 (0, 1/3, 2/3, 1) |
| Severe anxiety | Self-reported use of anxiolytic or sedative medications | Self-reported diagnosis of anxiety | No = 0; yes = 1 |
| Depression | Frequency of feeling that life is entirely hopeless in the past 2 weeks compared to usual (not at all, no more than usual, rather more than usual, much more than usual) | Frequency of depressed mood in last 2 weeks (not at all, several days, more than half the days, nearly every day) | Scaled on scale of 0-1 (0, 1/3, 2/3, 1) |
| Anhedonia | Frequency of being able to enjoy normal day-to-day activities in the past 2 weeks compared to usual (not at all, no more than usual, rather more than usual, much more than usual) | Frequency of little interest or pleasure in doing things in last 2 weeks (not at all, several days, more than half the days, nearly every day) | Scaled on scale of 0-1 (0, 1/3, 2/3, 1) |
| Fatigue | Frequency of feeling run down and out of sorts in the past 2 weeks compared to usual (not at all, no more than usual, rather more than usual, much more than usual) | Frequency of tiredness or lethargy in last 2 weeks (not at all, several days, more than half the days, nearly every day) | Scaled on scale of 0-1 (0, 1/3, 2/3, 1) |
| Sleep | Frequency of have trouble falling asleep at night OR waking up in the middle of the night in the past 2 weeks compared to usual (not at all, no more than usual, rather more than usual, much more than usual) OR use of insomnia medications | Frequency of having trouble falling asleep at night or do you wake up in the middle of the night (never/rarely, sometimes, usually) | Scaled on scale of 0-1 (NSHD: 0, 1/3, 2/3, 1; UKB: 0, 0.5, 1) and use of insomnia medications = 1 |
| Rheumatoid arthritis | Use of anti-rheumatic medications | Self-report of rheumatoid arthritis diagnosis | No = 0; yes = 1 |
| Gout | Use of gout medications | Self-report of gout diagnosis | No = 0; yes = 1 |
| Osteoporosis | Bone density t-score by DXA scan ≤2.5 at spine, femoral neck, or hip^5^ OR use of medications affecting bone metabolism | Self-report of osteoporosis diagnosis | No = 0; yes = 1 |
| Back pain and sciatica | Self-report of sciatica, lumbago or severe backache in the past 12 months | Self-report of sciatica diagnosis OR self-report of back pain interfering with usual activities in the past month | No = 0; yes = 1 |
| Body pain | Self-report of pain which interferes with normal work | Self-report of body pain interfering with usual activities | No = 0; yes = 1 |
| Head pain | Self-report of recently getting pains in their head | Self-report of head pain interfering with usual activities or headaches | No = 0; yes = 1 |
| Knee pain | Self-report of pain in and around knees on most days of the month for at least 3 months | Self-report of back pain interfering with usual activities in the past month | No = 0; yes = 1 |
| COPD | Cough or phlegm lasting 3 weeks or more in the past 3 years AND bringing up phlegm on most days for 3 months in the year | Self-reported chronic bronchitis/emphysema | No = 0; yes = 1 |
| Respiratory illness | Any chest illness, (e.g., bronchitis or pneumonia) which kept them off work or indoors for a week or more in the past 3 years | Self-report of pneumonia or bronchitis | No = 0; yes = 1 |
| Wheeze | Ever had wheezy or whistling sounds in the chest | Wheeze or whistling in the chest in last year | No = 0; yes = 1 |
| Glaucoma | Use of anti-glaucoma medication | Self-report of glaucoma diagnosis | No = 0; yes = 1 |
| Hearing | Average frequency of having difficulty hearing a normal conversation, a conversation in a noisy room, or over the phone in the past 12 months (rarely/never, sometimes, often, very often) | Self-report of difficulty with hearing | UKB: No = 0; yes = 1; NSHD: 0, 1/9, 2/9, 3/9, 4/9, 5/9, 6/9/, 7/9, 8/9, 1 |
| Vision/ cataracts | Average frequency of having difficulty with reading a newspaper, recognising a friend across the street, or reading signs at night (rarely/never, sometimes, often, very often) | Self-report of cataracts diagnosis | UKB: No = 0, yes = 1; NSHD: 0, 1/9, 2/9, 3/9, 4/9, 5/9, 6/9/, 7/9, 8/9, 1 |
| Allergy/ asthma/ eczema | Self-report of asthma or hay fever OR use of allergy medications OR use of eczema medications | Self-report of hay fever, allergic rhinitis, eczema or psoriasis | No = 0; yes = 1 |
| GERD/ indigestion/ heartburn | Use of anti-acid medications | Self-report of gastro-oesophageal reflux or gastric reflux OR use of ranitidine OR use of omeprazole | No = 0; yes = 1 |
| Constipation | Use of laxative medications | Use of laxative medications | No = 0; yes = 1 |

eTable 5. Descriptives of frailty index items

| **Item** | **NSHD** | | | **UKB** | | |
| --- | --- | --- | --- | --- | --- | --- |
|  | N (%) missing | CC Mean (SD) | MI Mean (SD) | N (%) missing | CC Mean (SD) | MI Mean (SD) |
| Cancer | - | 0.153 (0.36) | 0.153 (0.36) | 35 (0.37) | 0.089 (0.29) | 0.089 (0.29) |
| Anaemia | 28 (1.56) | 0.054 (0.23) | 0.058 (0.23) | 8 (0.08) | 0.013 (0.11) | 0.013 (0.11) |
| Angina | 6 (0.34) | 0.055 (0.23) | 0.055 (0.23) | 34 (0.36) | 0.033 (0.18) | 0.035 (0.18) |
| Chest pain | 9 (0.50) | 0.213 (0.41) | 0.206 (0.40) | 111 (1.18) | 0.155 (0.36) | 0.160 (0.37) |
| Diabetes | 4 (0.22) | 0.069 (0.25) | 0.072 (0.26) | 37 (0.39) | 0.049 (0.22) | 0.051 (0.22) |
| Myocardial infarction | 12 (0.67) | 0.036 (0.19) | 0.035 (0.18) | 33 (0.35) | 0.022 (0.15) | 0.022 (0.15) |
| High blood pressure | 5 (0.28) | 0.566 (0.50) | 0.570 (0.50) | 8 (0.08) | 0.650 (0.48) | 0.659 (0.47) |
| Raised cholesterol | - | 0.245 (0.43) | 0.244 (0.43) | 76 (0.81) | 0.165 (0.37) | 0.171 (0.38) |
| Stroke | 43 (2.40) | 0.017 (0.13) | 0.018 (0.13) | 33 (0.35) | 0.012 (0.11) | 0.014 (0.12) |
| Thyroid disorder | 141 (7.87) | 0.080 (0.27) | 0.086 (0.28) | 7 (0.07) | 0.052 (0.22) | 0.055 (0.23) |
| BMI | 3 (0.17) | 0.490 (0.38) | 0.490 (0.38) | 640 (6.79) | 0.462 (0.38) | 0.460 (0.38) |
| Waist to hip ratio | 6 (0.34) | 0.717 (0.45) | 0.721 (0.45) | 521 (5.53) | 0.238 (0.43) | 0.246 (0.43) |
| Recent weight lost | 6 (0.34) | 0.047 (0.21) | 0.047 (0.21) | 651 (6.91) | 0.158 (0.36) | 0.158 (0.37) |
| Epilepsy | 144 (8.04) | 0.024 (0.15) | 0.025 (0.16) | 8 (0.08) | 0.010 (0.10) | 0.010 (0.10) |
| General health | 146 (8.15) | 0.364 (0.23) | 0.369 (0.23) | 60 (0.64) | 0.371 (0.25) | 0.378 (0.25) |
| Polypharmacy | - | 0.188 (0.39) | 0.190 (0.39) | 4 (0.04) | 0.175 (0.38) | 0.183 (0.39) |
| Long-term illness | 4 (0.22) | 0.239 (0.43) | 0.240 (0.43) | 253 (2.68) | 0.320 (0.47) | 0.328 (0.47) |
| Falls | 11 (0.61) | 0.174 (0.38) | 0.174 (0.38) | 31 (0.33) | 0.190 (0.39) | 0.196 (0.40) |
| Fractures | 82 (4.58) | 0.078 (0.27) | 0.080 (0.27) | 67 (0.71) | 0.093 (0.29) | 0.095 (0.29) |
| Anxiety | 20 (1.12) | 0.134 (0.21) | 0.134 (0.21) | 872 (9.25) | 0.110 (0.21) | 0.112 (0.21) |
| Severe anxiety | - | 0.019 (0.14) | 0.020 (0.14) | 8 (0.08) | 0.013 (0.11) | 0.012 (0.11) |
| Depression | 20 (1.12) | 0.045 (0.13) | 0.047 (0.14) | 410 (4.35) | 0.101 (0.20) | 0.104 (0.21) |
| Anhedonia | 20 (1.12) | 0.354 (0.15) | 0.354 (0.16) | 858 (9.10) | 0.092 (0.20) | 0.096 (0.21) |
| Fatigue | 21 (1.17) | 0.226 (0.25) | 0.228 (0.25) | 298 (3.16) | 0.238 (0.28) | 0.241 (0.28) |
| Sleep | 21 (1.17) | 0.319 (0.23) | 0.321 (0.23) | 18 (0.19) | 0.514 (0.36) | 0.521 (0.36) |
| Rheumatoid arthritis | - | 0.014 (0.12) | 0.013 (0.12) | 8 (0.08) | 0.010 (0.10) | 0.011 (0.10) |
| Gout | - | 0.015 (0.12) | 0.014 (0.12) | 8 (0.08) | 0.014 (0.12) | 0.014 (0.12) |
| Osteoporosis | - | 0.097 (0.30) | 0.095 (0.29) | 8 (0.08) | 0.015 (0.12) | 0.016 (0.13) |
| Back pain and sciatica | 6 (0.34) | 0.298 (0.46) | 0.302 (0.46) | 35 (0.37) | 0.258 (0.44) | 0.262 (0.44) |
| Body pain | 160 (8.93) | 0.374 (0.48) | 0.383 (0.49) | 36 (0.38) | 0.016 (0.12) | 0.017 (0.13) |
| Head pain | 20 (1.12) | 0.274 (0.45) | 0.273 (0.45) | 36 (0.38) | 0.206 (0.40) | 0.207 (0.41) |
| Knee pain | 6 (0.34) | 0.222 (0.42) | 0.223 (0.42) | 36 (0.38) | 0.210 (0.41) | 0.215 (0.41) |
| Chronic bronchitis | 146 (8.15) | 0.061 (0.24) | 0.063 (0.24) | 32 (0.34) | 0.015 (0.12) | 0.017 (0.13) |
| Respiratory illness | 146 (8.15) | 0.113 (0.32) | 0.114 (0.32) | 8 (0.08) | 0.020 (0.14) | 0.020 (0.14) |
| Wheeze | 150 (8.38) | 0.196 (0.40) | 0.201 (0.40) | 223 (2.37) | 0.201 (0.40) | 0.205 (0.40) |
| Glaucoma | - | 0.014 (0.12) | 0.014 (0.12) | 599 (6.35) | 0.015 (0.12) | 0.031 (0.17) |
| Vision/ cataracts | 9 (0.50) | 0.130 (0.19) | 0.130 (0.19) | 483 (5.12) | 0.032 (0.18) | 0.045 (0.21) |
| Hearing | 9 (0.50) | 0.031 (0.09) | 0.032 (0.10) | 399 (4.23) | 0.256 (0.44) | 0.259 (0.44) |
| Allergy/ asthma/ eczema | 69 (3.85) | 0.231 (0.42) | 0.229 (0.42) | 31 (0.33) | 0.305 (0.46) | 0.300 (0.46) |
| GERD/ indigestion/ heartburn | - | 0.134 (0.34) | 0.133 (0.34) | 101 (1.07) | 0.093 (0.29) | 0.097 (0.30) |
| Constipation | - | 0.014 (0.12) | 0.016 (0.12) | 111 (1.18) | 0.027 (0.16) | 0.029 (0.17) |
| Note: All items on a scale of 0 to 1 where 0 represents the lowest possible deficit and 1 represents the highest possible deficit. All statistics for the valid sample. CC = complete case dataset, MI = multiple imputed datasets. | | | | | | |

eTable 6. Associations between serostatus and frailty index in minimally adjusted models

|  | **UKB** | | **NSHD** | | **Pooled** | | | | | | |
| --- | --- | --- | --- | --- | --- | --- | --- | --- | --- | --- | --- |
|  | Beta | 95% CI | Beta | 95% CI | Beta | 95% CI | P-value | P_FDR_ | UKB weighting | NSHD weighting | I^2^ |
| **Herpesviruses** |  |  |  |  |  |  |  |  |  |  |  |
| Herpes simplex virus-1 | 0.982 | 0.61, 1.35 | 1.642 | 0.75, 2.53 | 1.189 | 0.58, 1.80 | 1.2x10^-4^ | 1.7x10^-3^ | 69.0 | 31.0 | 46.1 |
| Herpes simplex virus-2 | 0.811 | 0.35, 1.27 | -1.235 | -2.97, 0.50 | -0.034 | -2.01, 1.94 | 0.973 | 0.981 | 58.7 | 41.3 | **80.0** |
| Varicella zoster virus | -0.019 | -0.66, 0.62 | 0.582 | -0.50, 1.66 | 0.139 | -0.41, 0.69 | 0.622 | 0.981 | 74.1 | 25.9 | 0 |
| Epstein-Barr virus | 0.979 | 0.22, 1.74 | 1.003 | -0.58, 2.58 | 0.985 | 0.30, 1.67 | 4.6x10^-3^ | 0.056 | 81.2 | 18.8 | 0 |
| Cytomegalovirus | 0.649 | 0.30, 1.00 | 0.520 | -0.34, 1.39 | 0.633 | 0.31, 0.96 | 1.4x10^-4^ | 1.8x10^-3^ | 85.9 | 14.1 | 0 |
| Human herpesvirus-6A | 0.175 | -0.23, 0.58 | 0.792 | -0.07, 1.66 | 0.357 | -0.19, 0.91 | 0.203 | 0.981 | 70.4 | 29.6 | 36.6 |
| Human herpesvirus-6B | -0.014 | -0.43, 0.40 | -0.124 | -0.99, 0.75 | -0.038 | -0.41, 0.34 | 0.844 | 0.981 | 81.4 | 18.6 | 0 |
| Human herpesvirus 7 | -0.120 | -0.88, 0.64 | -0.410 | -1.44, 0.62 | -0.227 | -0.84, 0.38 | 0.464 | 0.981 | 64.8 | 35.2 | 0 |
| Kaposi’s sarcoma-associated virus | 0.346 | -0.28, 0.97 | 5.257 | -4.66, 15.17 | 0.365 | -0.26, 0.99 | 0.250 | 0.981 | 99.6 | 0.4 | 0 |
| **Polyomaviruses** |  |  |  |  |  |  |  |  |  |  |  |
| BK virus | -0.423 | -1.22, 0.38 | -0.246 | -1.84, 1.35 | -0.391 | -1.11, 0.33 | 0.285 | 0.981 | 79.9 | 20.1 | 0 |
| JC virus | 0.189 | -0.15, 0.53 | -0.046 | -0.91, 0.82 | 0.153 | -0.17, 0.47 | 0.347 | 0.981 | 86.3 | 13.7 | 0 |
| Merkel Cell virus | -0.491 | -0.85, -0.13 | -1.278 | -2.17, -0.38 | -0.783 | -1.54, -0.03 | 0.043 | 0.472 | 63.6 | 36.4 | **62.5** |
| **Papillomaviruses** |  |  |  |  |  |  |  |  |  |  |  |
| Human Papillomavirus-16 | 0.221 | -0.60, 1.05 | -1.367 | -3.85, 1.12 | -0.108 | -1.37, 1.15 | 0.866 | 0.981 | 79.2 | 20.8 | 27.5 |
| Human Papillomavirus-18 | 0.107 | -0.94, 1.16 | -1.158 | -4.45, 2.13 | -0.012 | -1.01, 0.99 | 0.981 | 0.981 | 90.8 | 9.2 | 0 |
| **Bacteria/protozoa** |  |  |  |  |  |  |  |  |  |  |  |
| *T. gondii* | 0.959 | 0.58, 1.34 | 0.477 | -0.57, 1.53 | 0.901 | 0.55, 1.27 | 7.03x10^-7^ | 1.2x10^-5^ | 88.5 | 11.5 | 0 |
| *H. pylori* | 1.263 | 0.89, 1.63 | 1.215 | 0.05, 2.38 | 1.260 | 0.90, 1.61 | 2.19x10^-12^ | 4.2x10^-11^ | 90.9 | 9.1 | 0 |
| *C. trachomatis* | 0.833 | 0.41, 1.25 | 1.079 | -0.13, 2.28 | 0.860 | 0.45, 1.61 | 2.09x10^-5^ | 3.1x10^-4^ | 89.2 | 10.8 | 0 |
| **Pathogen burden indices** |  |  |  |  |  |  |  |  |  |  |  |
| Unweighted PBI | 0.542 | 0.37, 0.71 | 0.347 | -0.11, 0.80 | 0.517 | 0.36, 0.68 | 3.3x10^-10^ | 5.9x10^-9^ | 87.4 | 12.6 | 0 |
| CRP-weighted PBI | 0.750 | 0.58, 0.92 | 0.583 | 0.11, 1.06 | 0.732 | 0.57, 0.89 | 1.0x10^-18^ | 2.1x10^-17^ | 88.4 | 11.6 | 0 |
| WBC-weighted PBI | 0.689 | 0.52, 0.86 | 1.079 | 0.61, 1.54 | 0.828 | 0.45, 1.20 | 1.4x10^-5^ | 2.2x10^-4^ | 65.2 | 34.8 | **60.1** |
| Notes: Results from multiply imputed datasets. Minimally adjusted models control for sex, age and ethnicity (in UKB only). Significant heterogeneity (I^2^) indicated by bold font. BK: BK virus; CMV: cytomegalovirus; EBV: Epstein–Barr virus; HHV: human herpesvirus; HPV: human papillomavirus; HSV: herpes simplex virus; JC: John Cunningham virus; KSHV: Kaposi's sarcoma–associated herpesviruses; MCV: Merkel cell virus; PBI: pathogen burden index; CRP: C-reactive protein; WBC: White blood cell. | | | | | | | | | | | |

eTable 7. Associations between serostatus and frailty index in fully adjusted models

|  | **UKB** | | **NSHD** | | **Pooled** | | | | | | |
| --- | --- | --- | --- | --- | --- | --- | --- | --- | --- | --- | --- |
|  | Beta | 95% CI | Beta | 95% CI | Beta | 95% CI | P-value | P_FDR_ | UKB weighting | NSHD weighting | I^2^ |
| **Herpesviruses** |  |  |  |  |  |  |  |  |  |  |  |
| Herpes simplex virus-1 | 0.284 | -0.08, 0.65 | 0.639 | -0.24, 1.51 | 0.333 | 0.00, 0.67 | 0.053 | 0.794 | 85.1 | 14.9 | 0 |
| Herpes simplex virus-2 | 0.678 | 0.23, 1.13 | -1.441 | -3.12, 0.24 | -0.199 | -2.21, 1.81 | 0.846 | 0.846 | 57.8 | 42.2 | **81.9** |
| Varicella zoster virus | -0.012 | -0.63, 0.61 | 0.394 | -0.65, 1.44 | 0.095 | -0.44, 0.63 | 0.726 | 0.846 | 74.0 | 26.0 | 0 |
| Epstein-Barr virus | 0.649 | -0.09, 1.39 | 0.584 | -0.96, 2.13 | 0.629 | -0.04, 1.29 | 0.064 | 0.846 | 81.1 | 18.9 | 0 |
| Cytomegalovirus | 0.362 | 0.02, 0.70 | -0.097 | -0.93, 0.74 | 0.293 | -0.03, 0.62 | 0.076 | 0.846 | 85.1 | 14.9 | 1.7 |
| Human herpesvirus-6A | 0.116 | -0.28, 0.51 | 0.669 | -0.16, 1.50 | 0.252 | -0.20, 0.71 | 0.278 | 0.846 | 74.5 | 25.5 | 22.9 |
| Human herpesvirus-6B | -0.105 | -0.51, 0.30 | -0.144 | -0.98, 0.70 | -0.113 | -0.48, 0.25 | 0.541 | 0.846 | 81.2 | 18.8 | 0 |
| Human herpesvirus 7 | -0.115 | -0.85, 0.62 | -0.120 | -1.11, 0.87 | -0.119 | -0.71, 0.47 | 0.694 | 0.846 | 64.2 | 35.8 | 0 |
| Kaposi’s sarcoma-associated virus | 0.293 | -0.31, 0.90 | 4.14 | -6.68, 14.95 | 0.304 | -0.30, 0.91 | 0.323 | 0.846 | 99.7 | 0.3 | 0 |
| **Polyomaviruses** |  |  |  |  |  |  |  |  |  |  |  |
| BK virus | -0.196 | -0.97, 0.58 | -0.290 | -1.85, 1.27 | -0.217 | -0.91, 0.48 | 0.541 | 0.846 | 80.1 | 19.9 | 0 |
| JC virus | 0.305 | -0.03, 0.64 | -0.082 | -0.91, 0.74 | 0.249 | -0.06, 0.56 | 0.114 | 0.846 | 86.1 | 13.9 | 0 |
| Merkel Cell virus | -0.312 | -0.66, 0.04 | -1.073 | -1.93, -0.22 | -0.599 | -1.33, 0.14 | 0.110 | 0.846 | 63.0 | 37.0 | **63.5** |
| **Papillomaviruses** |  |  |  |  |  |  |  |  |  |  |  |
| Human Papillomavirus-16 | 0.095 | -0.71, 0.90 | -2.125 | -4.51, 0.26 | -0.718 | -2.81, 1.38 | 0.503 | 0.846 | 63.4 | 36.6 | **66.4** |
| Human Papillomavirus-18 | -0.079 | -1.10, 0.94 | -1.399 | -4.58, 1.78 | -0.201 | -1.17, 0.77 | 0.685 | 0.846 | 90.7 | 9.3 | 0 |
| **Bacteria/protozoa** |  |  |  |  |  |  |  |  |  |  |  |
| *T. gondii* | 0.810 | 0.44, 1.18 | 0.460 | -0.54, 1.46 | 0.767 | 0.42, 1.11 | 1.3x10^-5^ | 2.4x10^-4^ | 88.1 | 11.9 | 0 |
| *H. pylori* | 0.655 | 0.29, 1.02 | 0.348 | -0.79, 1.48 | 0.625 | 0.28, 0.97 | 3.9x10^-4^ | 6.6x10^-3^ | 90.8 | 9.2 | 0 |
| *C. trachomatis* | 0.576 | 0.17, 0.98 | 0.527 | -0.60, 1.65 | 0.571 | 0.19, 0.95 | 3.5x10^-3^ | 0.056 | 88.5 | 11.5 | 0 |
| **Pathogen burden indices** |  |  |  |  |  |  |  |  |  |  |  |
| Unweighted PBI | 0.340 | 0.15, 0.48 | 0.026 | -0.42, 0.48 | 0.245 | -0.05, 0.55 | 0.102 | 0.846 | 70.9 | 29.1 | 44.5 |
| CRP-weighted PBI | 0.435 | 0.27, 0.61 | 0.334 | -0.13, 0.80 | 0.424 | 0.26, 0.58 | 2.2x10^-7^ | 6.1x10^-6^ | 87.5 | 12.5 | 0 |
| WBC-weighted PBI | 0.383 | 0.21, 0.55 | 0.633 | 0.19, 1.08 | 0.413 | 0.25, 0.57 | 3.2x10^-5^ | 4.3x10^-4^ | 88.3 | 11.7 | 0 |
| Notes: Results from multiply imputed datasets. Fully adjusted models control for sex, age, ethnicity (in UKB only), income and educational attainment. Significant heterogeneity (I^2^) indicated by bold font. BK: BK virus; CMV: cytomegalovirus; EBV: Epstein–Barr virus; HHV: human herpesvirus; HPV: human papillomavirus; HSV: herpes simplex virus; JC: John Cunningham virus; KSHV: Kaposi's sarcoma–associated herpesviruses; MCV: Merkel cell virus; PBI: pathogen burden index; CRP: C-reactive protein; WBC: White blood cell. | | | | | | | | | | | |

eTable 8. Associations between serostatus and mortality in minimally adjusted models

|  | **UKB** | | **NSHD** | | **Pooled** | | | | | | |
| --- | --- | --- | --- | --- | --- | --- | --- | --- | --- | --- | --- |
|  | HR | 95% CI | HR | 95% CI | HR | 95% CI | P-value | P_FDR_ | UKB weighting | NSHD weighting | I^2^ |
| **Herpesviruses** |  |  |  |  |  |  |  |  |  |  |  |
| Herpes simplex virus-1 | 1.35 | 1.13, 1.61 | 0.91 | 0.71, 1.16 | 1.12 | 0.76, 1.65 | 0.576 | 0.934 | 52.4 | 47.6 | **85.0** |
| Herpes simplex virus-2 | 1.23 | 1.01, 1.49 | 1.26 | 0.83, 1.91 | 1.24 | 1.04, 1.47 | 0.018 | 0.313 | 82.3 | 17.7 | 0 |
| Varicella zoster virus | 0.84 | 0.63, 1.10 | 1.18 | 0.86, 1.62 | 0.99 | 0.70, 1.38 | 0.934 | 0.934 | 52.4 | 47.3 | **61.5** |
| Epstein-Barr virus | 1.61 | 1.09, 2.36 | 0.87 | 0.57, 1.31 | 1.19 | 0.65, 2.17 | 0.582 | 0.934 | 50.8 | 49.2 | **78.1** |
| Cytomegalovirus | 1.04 | 0.89, 1.21 | 0.90 | 0.71, 1.14 | 0.99 | 0.87, 1.13 | 0.914 | 0.934 | 70.0 | 30.0 | 0 |
| Human herpesvirus-6A | 1.03 | 0.86, 1.23 | 1.23 | 0.97, 1.55 | 1.11 | 0.93, 1.31 | 0.247 | 0.934 | 59.2 | 40.8 | 25.7 |
| Human herpesvirus-6B | 0.95 | 0.79, 1.13 | 1.20 | 0.95, 1.53 | 1.05 | 0.83, 1.34 | 0.661 | 0.934 | 55.3 | 44.7 | **61.2** |
| Human herpesvirus 7 | 0.85 | 0.64, 1.13 | 0.98 | 0.75, 1.27 | 0.92 | 0.76, 1.11 | 0.386 | 0.934 | 45.5 | 54.5 | 0 |
| Kaposi’s sarcoma-associated virus | 0.97 | 0.74, 1.28 | 1.20 | 0.19, 7.58 | 0.98 | 0.74, 1.28 | 0.861 | 0.934 | 97.8 | 2.2 | 0 |
| **Polyomaviruses** |  |  |  |  |  |  |  |  |  |  |  |
| BK virus | 0.67 | 0.05, 0.88 | 0.85 | 0.57, 1.26 | 0.72 | 0.57, 0.91 | 5.0x10^-3^ | 0.094 | 67.4 | 56.6 | 0 |
| JC virus | 1.24 | 1.06, 1.44 | 0.96 | 0.76, 1.21 | 1.11 | 0.87, 1.41 | 0.409 | 0.934 | 56.6 | 43.4 | **67.7** |
| Merkel Cell virus | 0.98 | 0.84, 1.15 | 1.00 | 0.79, 1.28 | 0.99 | 0.87, 1.13 | 0.884 | 0.934 | 70.7 | 29.3 | 0 |
| **Papillomaviruses** |  |  |  |  |  |  |  |  |  |  |  |
| Human Papillomavirus-16 | 1.66 | 1.18, 2.34 | 0.91 | 0.43, 1.91 | 1.35 | 0.77, 2.36 | 0.295 | 0.934 | 65.7 | 34.3 | **52.1** |
| Human Papillomavirus-18 | 1.50 | 0.97, 2.31 | 0.46 | 0.15, 1.41 | 0.93 | 0.30, 2.90 | 0.905 | 0.934 | 60.0 | 40.0 | **73.0** |
| **Bacteria/protozoa** |  |  |  |  |  |  |  |  |  |  |  |
| *T. gondii* | 1.18 | 1.01, 1.38 | 1.12 | 0.86, 1.46 | 1.17 | 1.02, 1.33 | 0.025 | 0.398 | 74.0 | 26.0 | 0 |
| *H. pylori* | 1.13 | 0.97, 1.32 | 1.21 | 0.90, 1.62 | 1.15 | 1.00, 1.31 | 0.051 | 0.710 | 78.7 | 21.3 | 0 |
| *C. trachomatis* | 1.22 | 1.01, 1.46 | 1.10 | 0.81, 1.51 | 1.19 | 1.01, 1.39 | 0.036 | 0.540 | 73.9 | 26.1 | 0 |
| **Pathogen burden indices** |  |  |  |  |  |  |  |  |  |  |  |
| Unweighted PBI | 1.12 | 1.04, 1.21 | 1.05 | 0.94, 1.19 | 1.11 | 1.04, 1.18 | 2.3x10^-3^ | 0.046 | 70.1 | 29.9 | 0 |
| CRP-weighted PBI | 1.16 | 1.07, 1.25 | 0.99 | 0.87, 1,12 | 1.08 | 0.92, 1.26 | 0.357 | 0.934 | 54.7 | 45.3 | **79.2** |
| WBC-weighted PBI | 1.13 | 1.05, 1.22 | 1.05 | 0.93, 1.19 | 1.11 | 1.03, 1.19 | 6.8x10^-3^ | 0.122 | 69.8 | 30.2 | 14.0 |
| Notes: Results from multiply imputed datasets. Minimally adjusted models control for sex and ethnicity (in UKB only). Significant heterogeneity (I^2^) indicated by bold font. BK: BK virus; CMV: cytomegalovirus; EBV: Epstein–Barr virus; HHV: human herpesvirus; HPV: human papillomavirus; HSV: herpes simplex virus; JC: John Cunningham virus; KSHV: Kaposi's sarcoma–associated herpesviruses; MCV: Merkel cell virus; PBI: pathogen burden index; CRP: C-reactive protein; WBC: White blood cell. | | | | | | | | | | | |

eTable 9. Associations between serostatus and mortality in fully adjusted models

|  | **UKB** | | **NSHD** | | **Pooled** | | | | | | |
| --- | --- | --- | --- | --- | --- | --- | --- | --- | --- | --- | --- |
|  | HR | 95% CI | HR | 95% CI | HR | 95% CI | P-value | P_FDR_ | UKB weighting | NSHD weighting | I^2^, |
| **Herpesviruses** |  |  |  |  |  |  |  |  |  |  |  |
| Herpes simplex virus-1 | 1.23 | 1.02, 1.47 | 0.82 | 0.63, 1.06 | 1.01 | 0.67, 1.51 | 0.967 | 0.977 | 52.6 | 47.4 | **84.9** |
| Herpes simplex virus-2 | 1.21 | 0.99, 1.47 | 1.22 | 0.79, 1.87 | 1.21 | 1.01, 1.44 | 0.034 | 0.654 | 83.0 | 17.0 | 0 |
| Varicella zoster virus | 0.83 | 0.63, 1.09 | 1.14 | 0.83, 1.57 | 0.96 | 0.70, 1.32 | 0.818 | 0.977 | 53.0 | 47.0 | **55.5** |
| Epstein-Barr virus | 1.51 | 1.03, 2.23 | 0.83 | 0.54, 1.27 | 1.12 | 0.62, 2.04 | 0.707 | 0.977 | 51.1 | 48.9 | **76.8** |
| Cytomegalovirus | 0.97 | 0.83, 1.13 | 0.84 | 0.66, 1.07 | 0.93 | 0.81, 1.06 | 0.266 | 0.977 | 71.1 | 28.9 | 0.3 |
| Human herpesvirus-6A | 1.02 | 0.85, 1.22 | 1.21 | 0.96, 1.54 | 1.09 | 0.93, 1.28 | 0.293 | 0.977 | 60.3 | 39.7 | 30.7 |
| Human herpesvirus-6B | 0.93 | 0.78, 1.12 | 1.19 | 0.93, 1.51 | 1.04 | 0.82, 1.32 | 0.748 | 0.977 | 55.5 | 44.5 | **61.1** |
| Human herpesvirus 7 | 0.84 | 0.63, 1.12 | 1.00 | 0.77, 1.31 | 0.93 | 0.76, 1.12 | 0.432 | 0.977 | 45.7 | 54.3 | 0 |
| Kaposi’s sarcoma-associated virus | 0.96 | 0.73, 1.27 | 0.98 | 0.17, 5.75 | 0.96 | 0.73, 1.26 | 0.782 | 0.977 | 97.7 | 2.3 | 0 |
| **Polyomaviruses** |  |  |  |  |  |  |  |  |  |  |  |
| BK virus | 0.68 | 0.51, 0.90 | 0.83 | 0.55, 1.24 | 0.73 | 0.58, 0.91 | 6.4x10^-3^ | 0.128 | 67.7 | 32.3 | 0 |
| JC virus | 1.26 | 1.09, 1.47 | 0.96 | 0.76, 1.21 | 1.12 | 0.85, 1.46 | 0.420 | 0.977 | 55.5 | 44.5 | **73.2** |
| Merkel Cell virus | 0.99 | 0.85, 1.15 | 1.04 | 0.81, 1.32 | 1.00 | 0.88, 1.14 | 0.977 | 0.977 | 70.8 | 29.2 | 0 |
| **Papillomaviruses** |  |  |  |  |  |  |  |  |  |  |  |
| Human Papillomavirus-16 | 1.63 | 1.16, 2.30 | 0.82 | 0.38, 1.76 | 1.26 | 0.66, 2.42 | 0.489 | 0.977 | 62.7 | 37.3 | **61.7** |
| Human Papillomavirus-18 | 1.47 | 0.95, 2.28 | 0.42 | 0.14, 1.29 | 0.88 | 0.26, 2.94 | 0.836 | 0.977 | 58.8 | 41.2 | **76.1** |
| **Bacteria/protozoa** |  |  |  |  |  |  |  |  |  |  |  |
| *T. gondii* | 1.14 | 0.98, 1.33 | 1.10 | 0.85, 1.44 | 1.13 | 0.99, 1.30 | 0.068 | 0.977 | 74.2 | 25.8 | 0 |
| *H. pylori* | 1.03 | 0.88, 1.20 | 1.11 | 0.82, 1.49 | 1.04 | 0.91, 1.20 | 0.555 | 0.977 | 78.9 | 21.1 | **67.1** |
| *C. trachomatis* | 1.17 | 0.97, 1.41 | 1.06 | 0.77, 1.44 | 1.14 | 0.97, 1.34 | 0.104 | 0.977 | 73.5 | 26.5 | 0 |
| **Pathogen burden indices** |  |  |  |  |  |  |  |  |  |  |  |
| Unweighted PBI | 1.09 | 1.04, 1.22 | 1.02 | 0.90, 1.15 | 1.06 | 1.00, 1.14 | 0.058 | 0.977 | 70.8 | 29.2 | 0 |
| CRP-weighted PBI | 1.10 | 1.02, 1.19 | 0.97 | 0.86, 1.10 | 1.04 | 0.92, 1.19 | 0.520 | 0.977 | 56.7 | 43.3 | **68.2** |
| WBC-weighted PBI | 1.08 | 1.00, 1.17 | 1.01 | 0.89, 1.14 | 1.06 | 0.99, 1.13 | 0.107 | 0.977 | 70.9 | 29.1 | 4.1 |
| Notes: Results from multiply imputed datasets. Fully adjusted models control for sex, ethnicity (in UKB only), income and educational attainment. Significant heterogeneity (I^2^) indicated by bold font. BK: BK virus; CMV: cytomegalovirus; EBV: Epstein–Barr virus; HHV: human herpesvirus; HPV: human papillomavirus; HSV: herpes simplex virus; JC: John Cunningham virus; KSHV: Kaposi's sarcoma–associated herpesviruses; MCV: Merkel cell virus; PBI: pathogen burden index; CRP: C-reactive protein; WBC: White blood cell. | | | | | | | | | | | |

eTable 10. Associations between serostatus and frailty index in minimally adjusted models using complete case datasets.

|  | **UKB** | | **NSHD** | | **Pooled** | | | | | |
| --- | --- | --- | --- | --- | --- | --- | --- | --- | --- | --- |
|  | Beta | 95% CI | Beta | 95% CI | Beta | 95% CI | P-value | UKB weighting | NSHD weighting | I^2^ |
| **Herpesviruses** |  |  |  |  |  |  |  |  |  |  |
| Herpes simplex virus-1 | 0.842 | 0.45, 1.24 | 1.693 | 0.53, 2.85 | 1.091 | 0.33, 1.85 | 5.2x10^-3^ | 71.0 | 29.0 | 47.0 |
| Herpes simplex virus-2 | 0.510 | 0.01, 1.01 | -1.041 | -3.06, 0.98 | 0.057 | -1.32, 1.44 | 0.935 | 70.8 | 29.2 | **53.1** |
| Varicella zoster virus | 0.103 | -0.58, 0.79 | 1.154 | -0.26, 2.57 | 0.438 | -0.52, 1.40 | 0.370 | 68.1 | 31.9 | 41.5 |
| Epstein-Barr virus | 0.713 | -0.11, 1.53 | -0.773 | -3.10, 1.56 | 0.383 | -0.83, 1.59 | 0.535 | 77.9 | 22.1 | 28.5 |
| Cytomegalovirus | 0.564 | 0.19, 0.94 | 0.479 | -0.65, 1.61 | 0.556 | 0.2,0 0.91 | 2.2x10^-3^ | 90.1 | 9.9 | 0 |
| Human herpesvirus-6A | 0.207 | -0.23, 0.64 | 0.663 | -0.47, 1.80 | 0.266 | -0.14, 0.67 | 0.198 | 12.7 | 87.3 | 0 |
| Human herpesvirus-6B | 0.005 | -0.44, 0.45 | 0.223 | -0.90, 1.35 | 0.034 | -0.38, 0.45 | 0.872 | 13.7 | 86.3 | 0 |
| Human herpesvirus 7 | 0.152 | -0.66, 0.97 | -0.263 | -1.61, 1.08 | 0.039 | -0.66, 0.74 | 0.912 | 27.0 | 73.0 | 0 |
| Kaposi’s sarcoma-associated virus | 0.243 | -0.42, 0.90 | 6.631 | -8.2, 21.46 | 0.256 | -0.41, 0.92 | 0.449 | 0.2 | 99.8 | 0 |
| **Polyomaviruses** |  |  |  |  |  |  |  |  |  |  |
| BK virus | 0.011 | -0.85, 0.87 | 0.110 | -1.90, 2.12 | 0.026 | -0.76, 0.82 | 0.949 | 84.5 | 15.5 | 0 |
| JC virus | 0.233 | -0.14, 0.60 | 0.534 | -0.58, 1.65 | 0.262 | -0.09, 0.61 | 0.141 | 90.2 | 9.8 | 0 |
| Merkel Cell virus | -0.503 | -0.89, -0.12 | -0.592 | -1.75, 0.56 | -0.512 | -0.88, -0.15 | 6.1x10^-3^ | 90.0 | 10.0 | 0 |
| **Papillomaviruses** |  |  |  |  |  |  |  |  |  |  |
| Human Papillomavirus-16 | 0.338 | -0.53, 1.21 | -0.672 | -4.08, 2.74 | 0.276 | -0.57, 1.12 | 0.523 | 93.8 | 6.2 | 0 |
| Human Papillomavirus-18 | 0.253 | -0.87, 1.37 | -1.214 | -5.58, 3.15 | 0.163 | -0.92, 1.25 | 0.769 | 93.8 | 6.2 | 0 |
| **Bacteria/protozoa** |  |  |  |  |  |  |  |  |  |  |
| *T.gondii* | 0.852 | 0.44, 1.26 | 0.588 | -0.85, 2.02 | 0.832 | 0.44, 1.22 | 3.1x10^-5^ | 92.5 | 7.5 | 0 |
| *H.pylori* | 1.204 | 0.81, 1.60 | 1.114 | -0.41, 2.63 | 1.198 | 0.81, 1.58 | 9.3x10^-10^ | 93.6 | 6.4 | 0 |
| *C.trachomatis* | 0.882 | 0.43, 1.33 | 1.526 | -0.08, 3.13 | 1.332 | 0.00, 2.67 | 2.6x10^-5^ | 92.7 | 7.3 | 0 |
| **Pathogen burden indices** |  |  |  |  |  |  |  |  |  |  |
| Unweighted PBI | 0.510 | 0.33, 0.70 | 0.624 | 0.03, 1.22 | 0.520 | 0.34, 0.70 | 7.9x10^-9^ | 91.3 | 8.7 | 0 |
| CRP-weighted PBI | 0.692 | 0.51, 0.88 | 0.247 | -0.41, 0.90 | 0.586 | 0.22, 0.96 | 1.9x10^-3^ | 76.2 | 23.8 | 38.5 |
| WBC-weighted PBI | 0.640 | 0.46, 0.82 | 0.942 | 0.35, 1.54 | 0.667 | 0.49, 0.84 | 1.1x10^-13^ | 91.2 | 8.8 | 0 |
| Notes: Minimally adjusted models control for sex, age and ethnicity (in UKB only). Significant heterogeneity (I^2^) indicated by bold font. BK: BK virus; CMV: cytomegalovirus; EBV: Epstein–Barr virus; HHV: human herpesvirus; HPV: human papillomavirus; HSV: herpes simplex virus; JC: John Cunningham virus; KSHV: Kaposi's sarcoma–associated herpesviruses; MCV: Merkel cell virus; PBI: pathogen burden index; CRP: C-reactive protein; WBC: White blood cell. | | | | | | | | | | |

eTable 11. Associations between serostatus and frailty index in fully adjusted models using complete case datasets.

|  | **UKB** | | **NSHD** | | **Pooled** | | | | | |
| --- | --- | --- | --- | --- | --- | --- | --- | --- | --- | --- |
|  | Beta | 95% CI | Beta | 95% CI | Beta | 95% CI | P-value | UKB weighting | NSHD weighting | I^2^ |
| **Herpesviruses** |  |  |  |  |  |  |  |  |  |  |
| Herpes simplex virus-1 | 0.188 | -0.20, 0.58 | 0.669 | -0.47, 1.81 | 0.238 | -0.13, 0.61 | 0.204 | 89.5 | 10.5 | 0 |
| Herpes simplex virus-2 | 0.430 | -0.05, 0.91 | -1.218 | -3.22, 0.78 | -0.097 | -1.60, 1.41 | 0.899 | 68.0 | 32.0 | **59.6** |
| Varicella zoster virus | 0.118 | -0.55, 0.78 | 0.913 | -0.43, 2.25 | 0.295 | -0.35, 0.94 | 0.373 | 77.7 | 22.3 | 8.1 |
| Epstein-Barr virus | 0.370 | -0.43, 1.17 | -1.069 | -3.24, 1.10 | 0.016 | -1.20, 1.23 | 0.979 | 75.5 | 24.5 | 33.1 |
| Cytomegalovirus | 0.329 | -0.03, 0.69 | -0.257 | -1.35, 0.83 | 0.270 | -0.07, 0.62 | 0.125 | 90.0 | 10.0 | 0 |
| Human herpesvirus-6A | 0.102 | -0.32, 0.52 | 0.566 | -0.52, 1.65 | 0.162 | -0.23, 0.55 | 0.417 | 87.0 | 13.0 | 0 |
| Human herpesvirus-6B | -0.097 | -0.53, 0.34 | -0.088 | -1.17, 0.99 | -0.096 | -0.50, 0.31 | 0.640 | 86.1 | 13.9 | 0 |
| Human herpesvirus 7 | 0.142 | -0.65, 0.93 | 0.136 | -1.14, 1.41 | 0.140 | -0.53, 0.81 | 0.683 | 72.1 | 27.9 | 0 |
| Kaposi’s sarcoma-associated virus | 0.177 | -0.46, 0.82 | 6.501 | -8.60, 21.60 | 0.189 | -0.45, 0.83 | 0.564 | 99.8 | 0.2 | 0 |
| **Polyomaviruses** |  |  |  |  |  |  |  |  |  |  |
| BK virus | 0.220 | -0.61, 1.05 | -0.077 | -2.00, 1.85 | 0.173 | -0.59, 0.94 | 0.658 | 84.2 | 15.8 | 0 |
| JC virus | 0.356 | 0.00, 0.71 | 0.420 | -0.63, 1.47 | 0.362 | 0.02, 0.70 | 0.035 | 89.7 | 10.3 | 0 |
| Merkel Cell virus | -0.358 | -0.73, 0.02 | -0.273 | -1.36, 0.81 | -0.349 | -0.70, 0.00 | 0.053 | 89.4 | 10.6 | 0 |
| **Papillomaviruses** |  |  |  |  |  |  |  |  |  |  |
| Human Papillomavirus-16 | 0.164 | -0.68, 1.01 | -1.143 | -4.03, 1.75 | 0.061 | -0.75, 0.87 | 0.884 | 92.1 | 7.9 | 0 |
| Human Papillomavirus-18 | 0.033 | -1.05, 1.12 | -0.899 | -5.20, 3.40 | -0.023 | -1.08, 1.03 | 0.966 | 94.0 | 6.0 | 0 |
| **Bacteria/protozoa** |  |  |  |  |  |  |  |  |  |  |
| *T.gondii* | 0.703 | 0.31, 1.10 | 0.604 | -0.77, 1.98 | 0.696 | 0.32, 1.08 | 3.3x10^-4^ | 92.4 | 7.6 | 0 |
| *H.pylori* | 0.633 | 0.24, 1.02 | 0.174 | -1.27, 1.62 | 0.602 | 0.23, 0.98 | 1.7x10^-3^ | 93.2 | 6.8 | 0 |
| *C.trachomatis* | 0.626 | 0.19, 1.06 | 0.957 | -0.50, 2.42 | 0.653 | 0.24, 1.07 | 2.2x10^-3^ | 91.8 | 8.2 | 0 |
| **Pathogen burden indices** |  |  |  |  |  |  |  |  |  |  |
| Unweighted PBI | 0.314 | 0.13, 0.49 | 0.273 | -0.32, 0.86 | 0.311 | 0.14, 0.48 | 4.2x10^-4^ | 91.4 | 8.6 | 0 |
| CRP-weighted PBI | 0.391 | 0.21, 0.57 | -0.030 | -0.67, 0.61 | 0.296 | -0.05, 0.64 | 0.091 | 77.5 | 22.5 | 35.1 |
| WBC-weighted PBI | 0.340 | 0.16, 0.52 | 0.387 | -0.18, 0.95 | 0.345 | 0.17, 0.52 | 9.4x10^-5^ | 90.6 | 9.4 | 0 |
| Notes: Fully adjusted models control for sex, age, ethnicity (in UKB only), income and educational attainment. Significant heterogeneity (I^2^) indicated by bold font. BK: BK virus; CMV: cytomegalovirus; EBV: Epstein–Barr virus; HHV: human herpesvirus; HPV: human papillomavirus; HSV: herpes simplex virus; JC: John Cunningham virus; KSHV: Kaposi's sarcoma–associated herpesviruses; MCV: Merkel cell virus; PBI: pathogen burden index; CRP: C-reactive protein; WBC: White blood cell. | | | | | | | | | | |

eTable 12. Associations between serostatus and mortality in minimally adjusted models using complete case datasets.

|  | **UKB** | | **NSHD** | | **Pooled** | | | | | |
| --- | --- | --- | --- | --- | --- | --- | --- | --- | --- | --- |
|  | HR | 95% CI | HR | 95% CI | HR | 95% CI | P-value | UKB weighting | NSHD weighting | I^2^ |
| **Herpesviruses** |  |  |  |  |  |  |  |  |  |  |
| Herpes simplex virus-1 | 1.38 | 1.14, 1.68 | 0.83 | 0.58, 1.19 | 1.08 | 0.66, 1.79 | 0.753 | 52.6 | 47.4 | **87.2** |
| Herpes simplex virus-2 | 1.09 | 0.87, 1.37 | 1.47 | 0.81, 2.66 | 1.17 | 0.92, 1.48 | 0.211 | 78.5 | 21.5 | 13.8 |
| Varicella zoster virus | 0.77 | 0.57, 1.04 | 0.73 | 0.48, 1.13 | 0.76 | 0.60, 0.95 | 0.015 | 57.0 | 43.0 | 0 |
| Epstein-Barr virus | 1.66 | 1.07, 2.57 | 0.65 | 0.37, 1.17 | 1.05 | 0.42, 2.62 | 0.913 | 51.0 | 49.0 | **86.6** |
| Cytomegalovirus | 1.05 | 0.89, 1.25 | 0.88 | 0.62, 1.25 | 1.00 | 0.85, 1.17 | 0.997 | 72.0 | 28.0 | 8.3 |
| Human herpesvirus-6A | 1.09 | 0.89, 1.33 | 1.30 | 0.92, 1.83 | 1.15 | 0.98, 1.36 | 0.090 | 66.1 | 33.9 | 0 |
| Human herpesvirus-6B | 1.00 | 0.82, 1.23 | 1.42 | 1.00, 2.02 | 1.17 | 0.83, 1.65 | 0.359 | 54.5 | 45.5 | **73.5** |
| Human herpesvirus 7 | 0.87 | 0.63, 1.20 | 1.05 | 0.71, 1.57 | 0.96 | 0.76, 1.20 | 0.700 | 49.6 | 50.4 | 0 |
| Kaposi’s sarcoma-associated virus | 1.00 | 0.75, 1.35 | 3.23 | 0.97, 10.74 | 1.44 | 0.50, 4.13 | 0.501 | 69.4 | 30.6 | **57.9** |
| **Polyomaviruses** |  |  |  |  |  |  |  |  |  |  |
| BK virus | 0.62 | 0.46, 0.83 | 1.00 | 0.53, 1.90 | 0.75 | 0.47, 1.19 | 0.223 | 59.7 | 40.3 | **60.3** |
| JC virus | 1.21 | 1.02, 1.43 | 0.87 | 0.62, 1.23 | 1.05 | 0.77, 1.44 | 0.759 | 56.6 | 43.4 | **72.7** |
| Merkel Cell virus | 1.02 | 0.86, 1.22 | 1.35 | 0.94, 1.95 | 1.14 | 0.87, 1.50 | 0.330 | 60.2 | 39.8 | **59.7** |
| **Papillomaviruses** |  |  |  |  |  |  |  |  |  |  |
| Human Papillomavirus-16 | 1.60 | 1.09, 2.34 | 1.14 | 0.43, 3.03 | 1.50 | 1.06, 2.11 | 0.021 | 80.9 | 19.1 | 0 |
| Human Papillomavirus-18 | 1.54 | 0.95, 2.50 | 0.13 | 0.02, 0.98 | 0.68 | 0.07, 6.72 | 0.744 | 67.2 | 32.8 | **63.5** |
| **Bacteria/protozoa** |  |  |  |  |  |  |  |  |  |  |
| *T. gondii* | 1.21 | 1.02, 1.44 | 1.26 | 0.86, 1.85 | 1.22 | 1.05, 1.42 | 8.6x10^-3^ | 76.9 | 23.1 | 0 |
| *H. pylori* | 1.13 | 0.95, 1.34 | 1.02 | 0.66, 1.57 | 1.10 | 0.95, 1.29 | 0.204 | 80.6 | 19.4 | 0 |
| *C. trachomatis* | 1.19 | 0.97, 1.47 | 1.20 | 0.76, 1.90 | 1.20 | 1.00, 1.43 | 0.052 | 76.4 | 23.6 | 0 |
| **Pathogen burden indices** |  |  |  |  |  |  |  |  |  |  |
| Unweighted PBI | 1.13 | 1.04, 1.23 | 1.08 | 0.90, 1.29 | 1.12 | 1.04, 1.21 | 2.6x10^-3^ | 74.5 | 25.5 | 0 |
| CRP-weighted PBI | 1.17 | 1.08, 1.27 | 0.84 | 0.71, 0.99 | 0.99 | 0.71, 1.38 | 0.967 | 51.5 | 48.5 | **93.9** |
| WBC-weighted PBI | 1.16 | 1.07, 1.26 | 0.97 | 0.80, 1.17 | 1.07 | 0.90, 1.28 | 0.038 | 55.9 | 44.1 | **76.7** |
| Notes: Minimally adjusted models control for sex and ethnicity (in UKB only). Significant heterogeneity (I^2^) indicated by bold font. BK: BK virus; CMV: cytomegalovirus; EBV: Epstein–Barr virus; HHV: human herpesvirus; HPV: human papillomavirus; HSV: herpes simplex virus; JC: John Cunningham virus; KSHV: Kaposi's sarcoma–associated herpesviruses; MCV: Merkel cell virus; PBI: pathogen burden index; CRP: C-reactive protein; WBC: White blood cell. | | | | | | | | | | |

eTable 13. Associations between serostatus and mortality in fully adjusted models using complete case datasets.

|  | **UKB** | | **NSHD** | | **Pooled** | | | | | |
| --- | --- | --- | --- | --- | --- | --- | --- | --- | --- | --- |
|  | HR | 95% CI | HR | 95% CI | HR | 95% CI | P-value | UKB weighting | NSHD weighting | I^2^ |
| **Herpesviruses** |  |  |  |  |  |  |  |  |  |  |
| Herpes simplex virus-1 | 1.27 | 1.04, 1.54 | 0.78 | 0.54, 1.12 | 1.00 | 0.62, 1.63 | 0.984 | 53.1 | 46.9 | **85.3** |
| Herpes simplex virus-2 | 1.08 | 0.87, 1.35 | 1.40 | 0.75, 2.60 | 1.13 | 0.92, 1.39 | 0.243 | 83.2 | 16.8 | 0 |
| Varicella zoster virus | 0.77 | 0.57, 1.03 | 0.70 | 0.46, 1.09 | 0.74 | 0.59, 0.93 | 8.9x10^-3^ | 57.1 | 42.9 | 0 |
| Epstein-Barr virus | 1.54 | 0.99, 2.38 | 0.65 | 0.36, 1.16 | 1.01 | 0.43, 2.35 | 0.984 | 51.3 | 48.7 | **84.1** |
| Cytomegalovirus | 0.99 | 0.84, 1.18 | 0.84 | 0.59, 1.21 | 0.95 | 0.82, 1.10 | 0.507 | 74.3 | 25.7 | 0 |
| Human herpesvirus-6A | 1.07 | 0.87, 1.31 | 1.31 | 0.93, 1.85 | 1.15 | 0.96, 1.39 | 0.138 | 63.4 | 36.6 | 17.9 |
| Human herpesvirus-6B | 0.98 | 0.80, 1.20 | 1.35 | 0.95, 1.93 | 1.13 | 0.83, 1.55 | 0.433 | 55.6 | 44.4 | **68.0** |
| Human herpesvirus 7 | 0.85 | 0.61, 1.17 | 1.06 | 0.71, 1.60 | 0.95 | 0.76, 1.19 | 0.655 | 50.1 | 49.9 | 0 |
| Kaposi’s sarcoma-associated virus | 0.99 | 0.73, 1.33 | 2.31 | 0.89, 6.03 | 1.10 | 0.63, 1.90 | 0.740 | 87.6 | 12.4 | 18.4 |
| **Polyomaviruses** |  |  |  |  |  |  |  |  |  |  |
| BK virus | 0.62 | 0.46, 0.84 | 0.96 | 0.50, 1.84 | 0.73 | 0.49, 1.10 | 0.132 | 62.5 | 37.5 | 49.4 |
| JC virus | 1.24 | 1.05, 1.47 | 0.86 | 0.61, 1.22 | 1.05 | 0.74, 1.50 | 0.770 | 55.4 | 44.6 | **77.9** |
| Merkel Cell virus | 1.02 | 0.86, 1.21 | 1.40 | 0.96, 2.03 | 1.17 | 0.86, 1.58 | 0.329 | 58.1 | 41.9 | **68.3** |
| **Papillomaviruses** |  |  |  |  |  |  |  |  |  |  |
| Human Papillomavirus-16 | 1.54 | 1.05, 2.26 | 0.98 | 0.38, 2.51 | 1.41 | 0.99, 2.01 | 0.059 | 80.2 | 19.8 | 3.0 |
| Human Papillomavirus-18 | 1.53 | 0.94, 2.49 | 0.14 | 0.02, 1.06 | 0.72 | 0.08, 6.39 | 0.768 | 68.5 | 31.5 | **60.8** |
| **Bacteria/protozoa** |  |  |  |  |  |  |  |  |  |  |
| *T. gondii* | 1.17 | 0.98, 1.39 | 1.18 | 0.81, 1.74 | 1.17 | 1.01, 1.36 | 0.039 | 77.0 | 23.0 | 0 |
| *H. pylori* | 1.03 | 0.87, 1.22 | 0.95 | 0.61, 1.49 | 1.01 | 0.08, 6.39 | 0.768 | 68.5 | 31.5 | 0 |
| *C. trachomatis* | 1.15 | 0.94, 1.42 | 1.16 | 0.74, 1.82 | 1.15 | 0.96, 1.38 | 0.124 | 76.7 | 23.3 | 0 |
| **Pathogen burden indices** |  |  |  |  |  |  |  |  |  |  |
| Unweighted PBI | 1.09 | 1.00, 1.19 | 1.04 | 0.87, 1.25 | 1.08 | 1.00, 1.16 | 0.046 | 75.3 | 24.7 | 0 |
| CRP-weighted PBI | 1.12 | 1.02, 1.22 | 0.84 | 0.71, 0.99 | 0.97 | 0.73, 1.29 | 0.841 | 52.0 | 48.0 | **91.4** |
| WBC-weighted PBI | 1.10 | 1.01, 1.20 | 0.94 | 0.78, 1.14 | 1.03 | 0.89, 1.20 | 0.687 | 57.6 | 42.4 | **69.7** |
| Notes: Fully adjusted models control for sex, ethnicity (in UKB only), income and educational attainment. Significant heterogeneity (I^2^) indicated by bold font. BK: BK virus; CMV: cytomegalovirus; EBV: Epstein–Barr virus; HHV: human herpesvirus; HPV: human papillomavirus; HSV: herpes simplex virus; JC: John Cunningham virus; KSHV: Kaposi's sarcoma–associated herpesviruses; MCV: Merkel cell virus; PBI: pathogen burden index; CRP: C-reactive protein; WBC: White blood cell. | | | | | | | | | | |
